# Semaglutide cardiovascular outcomes align more closely with attained dose than achieved weight loss

**DOI:** 10.64898/2026.04.02.26350077

**Authors:** Karthik Murugadoss, A.J. Venkatakrishnan, Christopher Gregg, Venky Soundararajan

## Abstract

Semaglutide is often optimized for weight loss, but whether longer-term cardiovascular benefit tracks achieved weight loss or therapeutic exposure levels remains unclear. Using a federated deidentified U.S. electronic health record network of 29 million patients, including 505,874 semaglutide-treated individuals, we leveraged multimodal AI technologies to analyze 47,199 patients with baseline cardiovascular disease. We quantified dose escalation and weight change during the 0–2-year period after semaglutide initiation (landmark period) and assessed cardiovascular outcomes during the 2–4-year period (post-landmark). In propensity-matched comparisons during the landmark period, semaglutide was associated with lower cardiovascular events than metformin, DPP-4 and SGLT2 inhibitors. Higher maximum semaglutide dose was associated with greater weight loss during the landmark period (3.15% additional weight loss per 1 mg increase; r=−0.97, P<0.001), and lower post-landmark risk of all-cause mortality (RR 0.42, p<0.001), composite cardiovascular events (death, myocardial infarction, or stroke; RR 0.51, p<0.001), cerebrovascular disease (RR 0.50, p<0.001), heart failure (RR 0.55, p<0.001), and valvular/rheumatic heart disease (RR 0.71, p=0.025). In contrast, greater achieved weight loss during the landmark period did not show a consistent monotonic association with lower post-landmark cardiovascular risk (All-cause mortality p-value=0.14, composite cardiovascular endpoint p-value=0.55). Integrating insights from a single cell GLP1R expression atlas was used to infer how semaglutide pharmacology may tie into heart-specific signaling, beyond what is reflected by body-weight reduction alone. The strongest prevalence-weighted GLP1R signal was observed in the pancreas, followed by the heart, where GLP1R engagement potential (GEP) was considerable across cardiomyocyte, cardiac endothelial, and rarer immune cell populations. Together, semaglutide cardiovascular benefit appears organized more by maximum dose attained than by achieved weight-loss magnitude, setting the stage for beyond-obesity trial designs that integrate whole-body spatial intelligence.

## Introduction

Cardiovascular disease is among the most consequential afterlives of excess adiposity, with myocardial injury, vascular dysfunction, and heart failure causing premature death across millions of patients worldwide^1^. In this landscape, semaglutide has emerged not merely as a drug that lowers body weight, but as a therapy that appears to reshape the clinical terrain of cardiometabolic disease itself. Randomized trials have shown that it can produce substantial and sustained weight reduction while also lowering major cardiovascular event risk in high-risk populations with type 2 diabetes and/orobesity^2–4^. Yet, despite its widening therapeutic reach, semaglutide is still most often understood by patients through the lens of a single visible metric: the magnitude of weight lost.

It is increasingly appreciated by physicians that this lens may be too narrow to capture the full biology of the drug. An expanding body of human evidence now suggests that semaglutide influences domains that extend beyond weight loss alone (**Table S1**)^5–7^, including major adverse cardiovascular events (SELECT), symptoms and physical limitations in obesity-related heart failure with preserved ejection fraction (STEP-HFpEF), kidney outcomes in type 2 diabetes with chronic kidney disease (FLOW), cardiovascular outcomes with oral semaglutide (SOUL), and walking capacity in symptomatic peripheral artery disease (STRIDE)^8–10^. The recent trial of high-dose semaglutide 7.2 mg provides an additional dose–response benchmark beyond the standard 0.25-2.4 mg range (STEP UP)^7^. Together, these findings raise the possibility that body-weight reduction, although important, is only one visible manifestation of a broader therapeutic biology.

This raises a question that is at once practical and mechanistic. In routine care, are dose, weight loss and cardiovascular benefit aligned along a single axis, or does semaglutide exert partially separable cardiovascular effects that are not fully captured by weight-loss magnitude? This question is especially relevant now that both treatment guidelines and commercial development strategies are increasingly organized around organ outcomes and comorbidity reduction, not weight loss alone^4,6^. Indeed, weight loss is a composite physiological readout, shaped not only by dose, but by treatment persistence, tolerability, metabolic context, baseline illness and individual biological variability. It is therefore possible that achieved weight reduction, although clinically salient, may serve as an incomplete surrogate for the signaling pathways most relevant to cardiovascular outcomes. Resolving this distinction is important not only for interpreting semaglutide response, but also for how cardiometabolic therapies are compared, optimized, and advanced beyond obesity alone.

Here, we addressed this question using deidentified longitudinal clinical data from the nSights Federated EHR Network, contextualized through a whole-body spatial intelligence framework^11–14^. As summarized in **Fig. 1**, the study couples a landmark clinical design and cohort-definition strategy with a broader conceptual framework linking therapeutic exposure, systems pharmacology, and diagnostics. Our findings position semaglutide as a cardiometabolic therapy whose clinical value may be organized along axes not captured by body-weight reduction alone and may be overlooked by conventional endpoints.

**Fig. 1.**
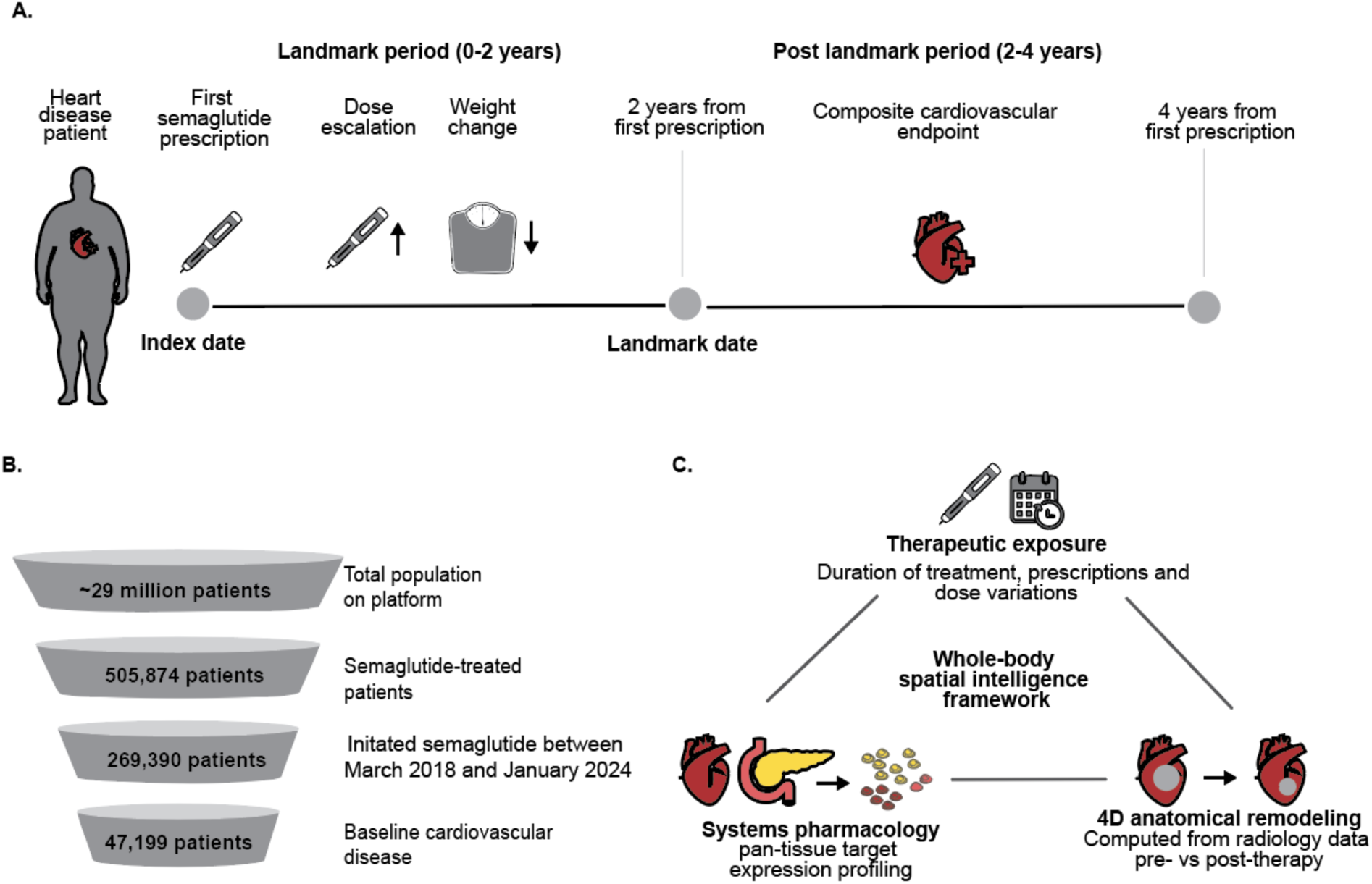
Study design and timeline for the semaglutide cardioprotective landmark analysis. (A) Patients initiating semaglutide between March 2018 and January 2024 were identified across the nSights Federated EHR Network. The index date was defined as the date of first semaglutide prescription. An incident user design was employed, requiring a one-year pre-index medication washout period to exclude patients with prior exposure to comparator medications. Baseline cardiovascular (CV) burden was ascertained from the entire available electronic health record (EHR) history prior to the index date, requiring ≥3 distinct ICD code dates per condition. Baseline body mass index (BMI) and body weight were captured as the nearest recorded value within windows of −365 to +14 days and −90 to +14 days relative to the index date, respectively. A two-year landmark date was defined for each patient, and only patients with documented follow-up through this date were included in the primary analysis. Dose escalation and weight loss were quantified over the landmark period (0–2 years). Post-landmark cardiovascular outcomes: all-cause mortality, composite cardiovascular events, and incident CV conditions, were assessed from the landmark date through the data cutoff of January 2026. (B) Cohort funnel showing the inclusion of patients based on semaglutide treatment and prior cardiovascular disease. (C) Conceptual framework for whole-body spatial intelligence connecting therapeutic exposure, systems pharmacology and 4D anatomical changes captured using radiological data.

## Results

### Higher attained semaglutide dose was associated with greater weight loss during the landmark period

Baseline demographic and clinical characteristics of patients stratified by maximum semaglutide dose are summarized in **Table S2**. Across the first two years after semaglutide initiation, attained dose and achieved weight loss were closely correlated (**Fig. 2**). Mean maximum weight loss increased from approximately 8% at 0.25 mg to 15% at 2.4 mg, corresponding to an estimated 3.15% greater maximum weight loss per 1 mg increase in attained dose (Pearson R = −0.97, P < 0.001).

**Fig. 2.**
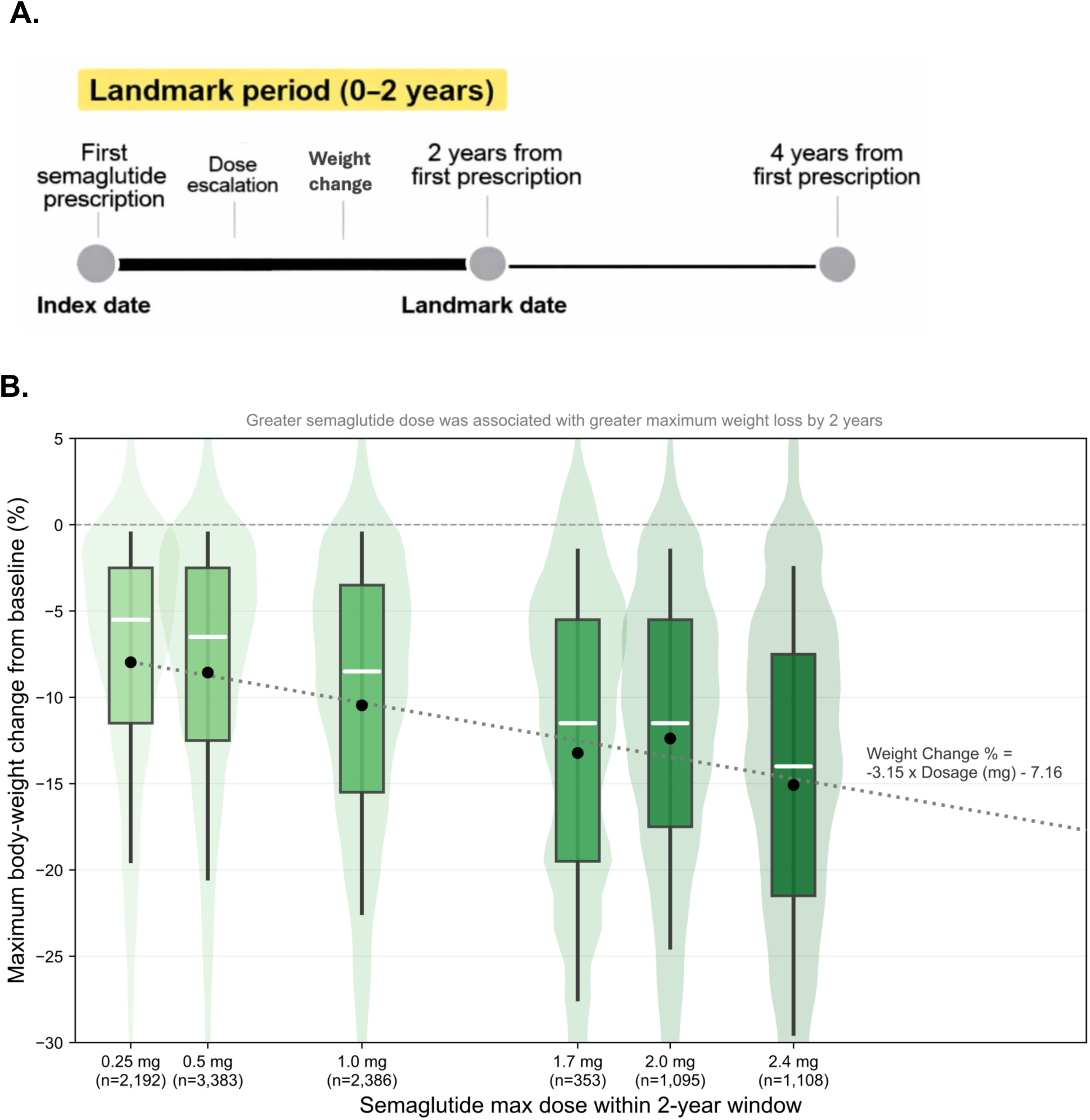
Relationship between semaglutide maximum dose and maximum weight loss through the 2-year landmark date. **(A)** Study design highlighting the 0-2 year landmark period where the current analysis is focused. **(B)** Boxplots display the distribution of maximum percent body-weight change achieved before the 2-year landmark according to the maximum semaglutide dose reached within the landmark window. Negative values indicate weight loss. Center lines indicate medians, boxes indicate interquartile ranges, whiskers indicate the spread of the data, and black dots indicate group means. The dotted line shows the fitted linear trend between dose and weight change, with the corresponding regression equation shown on the plot. Sample sizes are indicated below each dose category. Baseline demographic and clinical characteristics of patients stratified by maximum semaglutide dose are summarized in **Table S2**.

The same gradient emerged when patients were stratified by the maximum body-weight reduction achieved during the landmark period. Demographically, when comparing the <5% weight-loss and >25% weight-loss strata, mean age decreased from 58.2 to 56.2 years, female representation increased from 61.8% to 80.9%, baseline BMI rose modestly from 35.2 to 36.9 kg/m^2^, baseline weight rose from 103.1 kg to 105.6 kg, and baseline HbA1c fell from 6.6 to 6.2. Regarding the association between dose and weight loss, the proportion reaching the highest dose category increased from 20.9% to 49.6%, and median semaglutide prescription counts during the first 2 years increased from 1 [IQR 1 to 2] to 5 [IQR 2 to 11] (**Table 1**). Weight loss, therefore, seemed to reflect a combination of cumulative treatment exposure and baseline phenotype.

**Table 1.** Baseline and treatment characteristics across weight-loss categories among semaglutide-treated patients. Weight-loss categories were defined according to the maximum percentage reduction in body weight achieved within each landmark window. Baseline BMI and baseline weight are presented as mean (SD); dose intensity is summarized as the proportion reaching the highest dose category and prescription counts are summarized as median [interquartile range].

| Weight Loss Category | Number of unique patients | Age, Mean (SD) | Female, % | Baseline BMI, mean (SD) | Baseline Weight, kg mean (SD) | T2DM Prevalence at Baseline, % | Baseline HBA1C, Mean (SD) | Max Dose, 2-year n (%) | Prescription, 2-year median (IQR) |
| --- | --- | --- | --- | --- | --- | --- | --- | --- | --- |
| <5% | 4,050 | 58.2 (12.6) | 61.8 | 35.2 (6.4) | 103.1 (25.6) | 28.3 | 6.6 (1.7) | 847 (20.9%) | 1 [1, 2] |
| 5-10% | 2,805 | 59.8 (12.1) | 62.6 | 35.4 (6.0) | 102.6 (23.1) | 30.1 | 6.5 (1.6) | 763 (27.2%) | 2 [1, 5] |
| 10-15% | 1,943 | 60.0 (11.9) | 66.9 | 35.4 (5.8) | 101.5 (22.3) | 30.6 | 6.5 (1.7) | 655 (33.7%) | 2 [1, 7] |
| 15-20% | 1,223 | 58.4 (12.8) | 73.7 | 35.5 (5.5) | 101.0 (21.8) | 27.5 | 6.3 (1.4) | 504 (41.2%) | 4 [1, 9] |
| 20-25% | 707 | 57.9 (11.6) | 76.9 | 35.5 (5.2) | 100.8 (21.3) | 25.7 | 6.2 (1.3) | 325 (46.0%) | 5 [1, 10] |
| >25% | 884 | 56.2 (12.1) | 80.9 | 36.9 (5.7) | 105.6 (23.6) | 25.8 | 6.2 (1.4) | 438 (49.6%) | 5 [2, 11] |

### Higher attained dose during the landmark period was associated with lower post-landmark cardiovascular risk

When the post-landmark period was examined through the lens of dose attained during the first two years, patients who reached high-dose semaglutide by the landmark (≥1.7 mg; n = 3,794) had lower subsequent risk in the 2-4 years (“post-landmark”) period than those who remained in the low-dose range (0.25 to 1.0 mg; n=8,725) across several major outcomes (**Fig. 3**). High attained dose was associated with lower: all-cause mortality (RR = 0.42, P<0.001), risk of the composite cardiovascular endpoint (RR = 0.51, P < 0.001), incident cerebrovascular disease (RR = 0.50, P < 0.001), incident heart failure (RR = 0.55, P < 0.001) and incident valvular or rheumatic heart disease (RR = 0.71, P = 0.025) (see **Methods**).

**Fig. 3.**
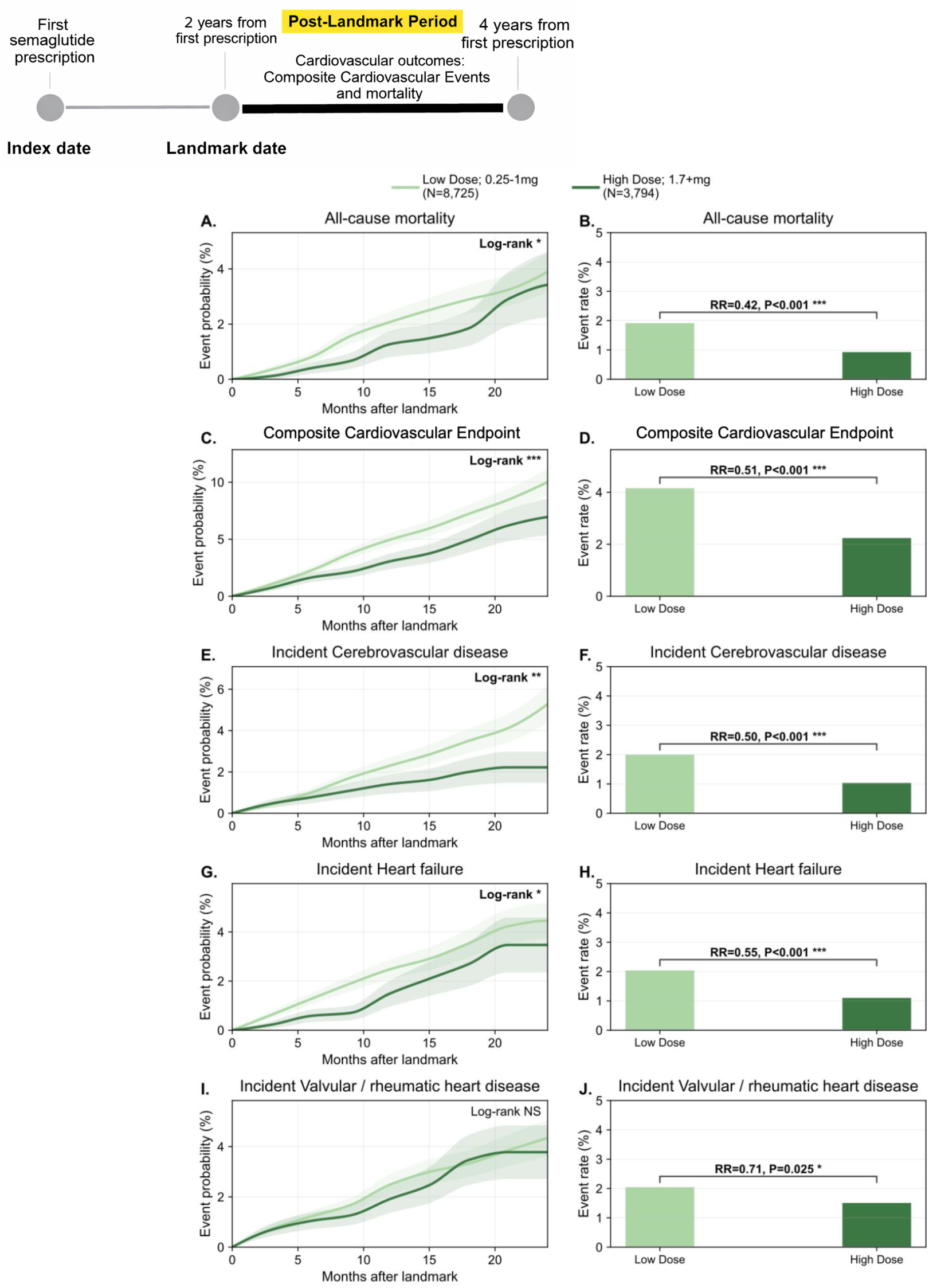
Higher maximum semaglutide dose was associated with lower post-landmark cardiovascular risk. Patients were grouped by the maximum semaglutide dose reached by the 2-year landmark as low dose (0.25–1.0 mg; *n* = 8,725) or high dose (≥1.7 mg; *n* = 3,794). Left-column panels show cumulative event curves after the landmark and right-column panels show corresponding post-landmark event rates for **(A-B)** all-cause mortality, **(C-D)** composite cardiovascular events, **(E-F)** incident cerebrovascular disease, **(G-H)** heart failure, and **(I-J)** incident valvular/rheumatic heart disease. Relative risks and nominal *P* values are shown on the bar plots. Higher dose semaglutide was associated with lower risk of all-cause mortality, composite cardiovascular events, incident cerebrovascular disease, incident heart failure, and valvular/rheumatic heart disease.

For all-cause mortality, the composite cardiovascular endpoint, cerebrovascular disease and heart failure, time-to-event analyses showed divergence that emerged within the first 5-10 months post-landmark and persisted through 24 months of follow-up (log-rank p<0.05 for all analyses) (**Fig. 3**). Cumulative event curves did not significantly differ for incident valvular or rheumatic heart disease. Ischemic heart disease, hypertension, arrhythmias or conduction disorders, peripheral vascular disease or atherosclerosis, cardiomyopathy, aortic disease and venous thromboembolism did not show clear dose-stratified differences in the post-landmark 2–4-year period (**Fig. S1**). Analysis anchored on the time of first attainment of maximum dose yielded directionally concordant results, again showing lower risk across the same cardiovascular outcomes in the higher-dose group **(Fig. S2**).

Dose attainment also correlated with treatment intensity beyond the landmark. During the first two years, median semaglutide prescription counts were 5 [IQR 1 to 9] in the high-dose group versus 1 [IQR 1 to 2] in the low-dose group (**Table S3**). In the post-landmark period (2-4 years upon initiating semaglutide), prescribing attenuated in both groups, but remained modestly higher in the high-dose group (median 1 [IQR 0 to 2] versus 0 [IQR 0 to 2]). However, larger negative pre-to-post-landmark changes in prescription counts were observed in the high-dose group (−4 [IQR −9 to −1] relative to the low-dose group −1 [IQR −2 to −1]).

### Higher weight loss during the landmark period was associated with post-landmark glycemic and systolic blood pressure control, but not later cardiovascular risk

When the same shared landmark was viewed through the lens of weight change rather than attained dose, the metabolic signals were as expected, but the cardiovascular signal was not.

Greater maximum weight loss during the first two years was accompanied by progressively more favorable metabolic-surrogate measures at and after the landmark. Post-landmark HbA1c fell from approximately 6.4% in the <5% weight-loss group to 5.6% in the >25% group, whereas patient-specific HbA1c change from the pre-treatment baseline deepened from −0.2% to −0.8% (one-way ANOVA P < 0.001; **Fig. 4a-b**). Post-landmark systolic blood pressure similarly declined from about 133 mmHg to 126 mmHg, with change from baseline shifting from roughly +2.0 to −3.5 mmHg (both P < 0.001; **Fig. 4c-d**). Post-landmark diastolic blood pressure declined more modestly, from about 78.0 to 75.4 mmHg, with progressively more negative change from baseline across strata (P = 0.002 for change from baseline). By conventional metabolic readouts, larger weight loss behaved exactly as a stronger systemic response should.

**Fig. 4.**
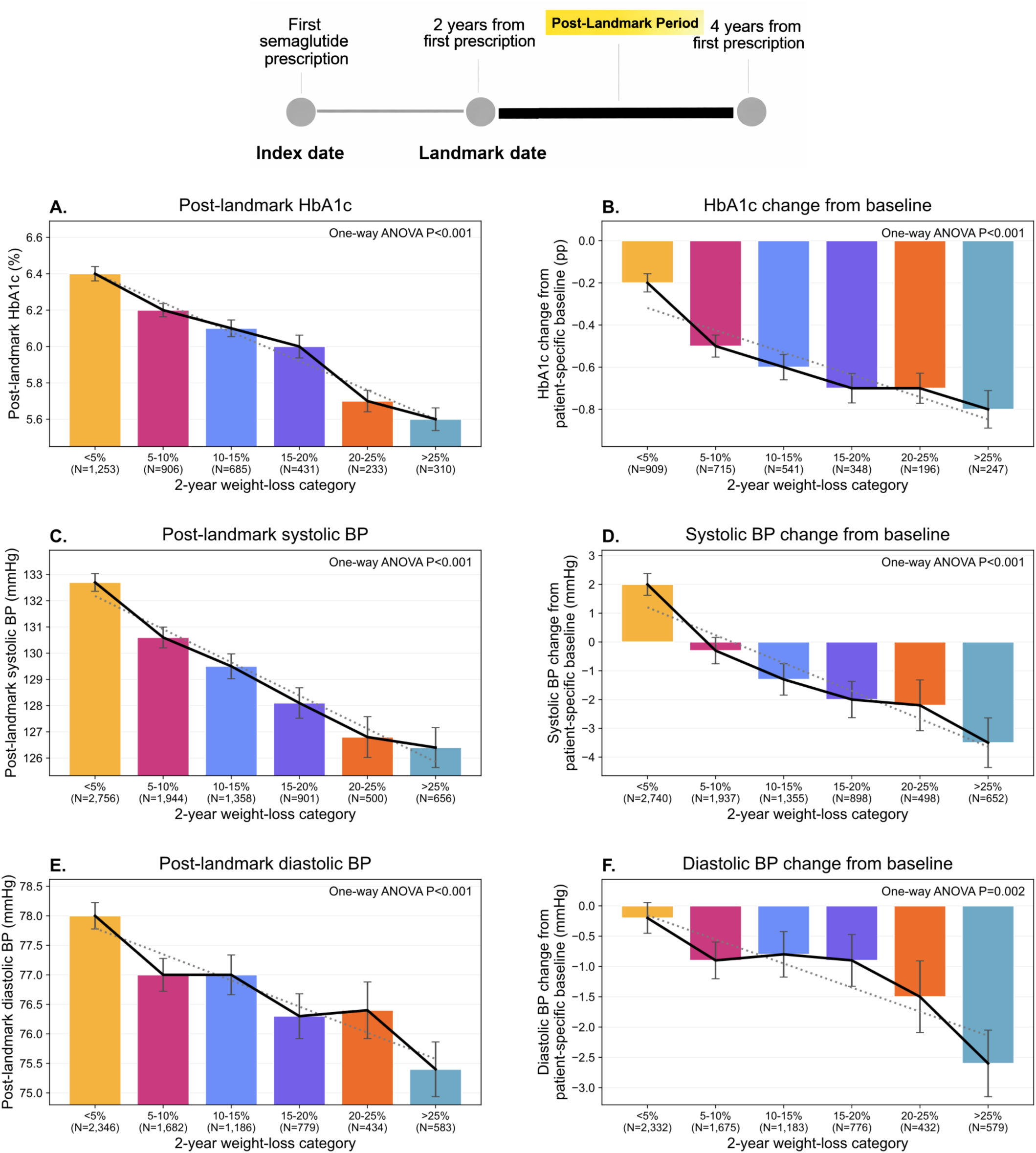
Semaglutide 2-year max weight-loss landmark analysis of HbA1c and blood pressure in the all cardiovascular conditions subgroup. Patients were categorized according to the maximum percentage reduction in body weight achieved before the 2-year landmark. Panels A and B show post-landmark HbA1c and change in HbA1c from patient-specific baseline, respectively, across weight-loss categories. Panels C and D show post-landmark systolic blood pressure and change from patient-specific baseline, and panels E and F show post-landmark diastolic blood pressure and change from patient-specific baseline. Bars indicate group means with standard error bars, dotted gray lines indicate fitted trends, solid black lines connect observed group means, and one-way ANOVA P values are shown.

Using the same 0 to 2-year weight-loss strata and the same 2-year landmark, post-landmark all-cause mortality, the composite cardiovascular endpoint, incident cerebrovascular disease, incident heart failure and incident valvular or rheumatic heart disease did not show a consistent monotonic gradient (**Fig. 5**). Global comparisons were not significant for all-cause mortality (P = 0.144), the composite cardiovascular endpoint (P = 0.547), incident cerebrovascular disease (P = 0.249), incident heart failure (P = 0.887) or incident valvular or rheumatic heart disease (P = 0.431). Among the additional cardiovascular outcomes, only incident peripheral vascular disease or atherosclerosis reached nominal significance across weight-loss strata (global P = 0.033), whereas incident hypertension showed only a suggestive downward trend with greater weight loss (global P = 0.107); the remaining outcomes were not significant (ischemic heart disease, arrhythmias, cardiomyopathy, aortic disease, and venous thromboembolism; **Fig. S3**). Taken together, these findings suggest that greater weight reduction during the 0-2 years landmark period did not necessarily translate into proportionally better cardiovascular outcomes in the 2-4 years post-landmark.

**Fig. 5.**
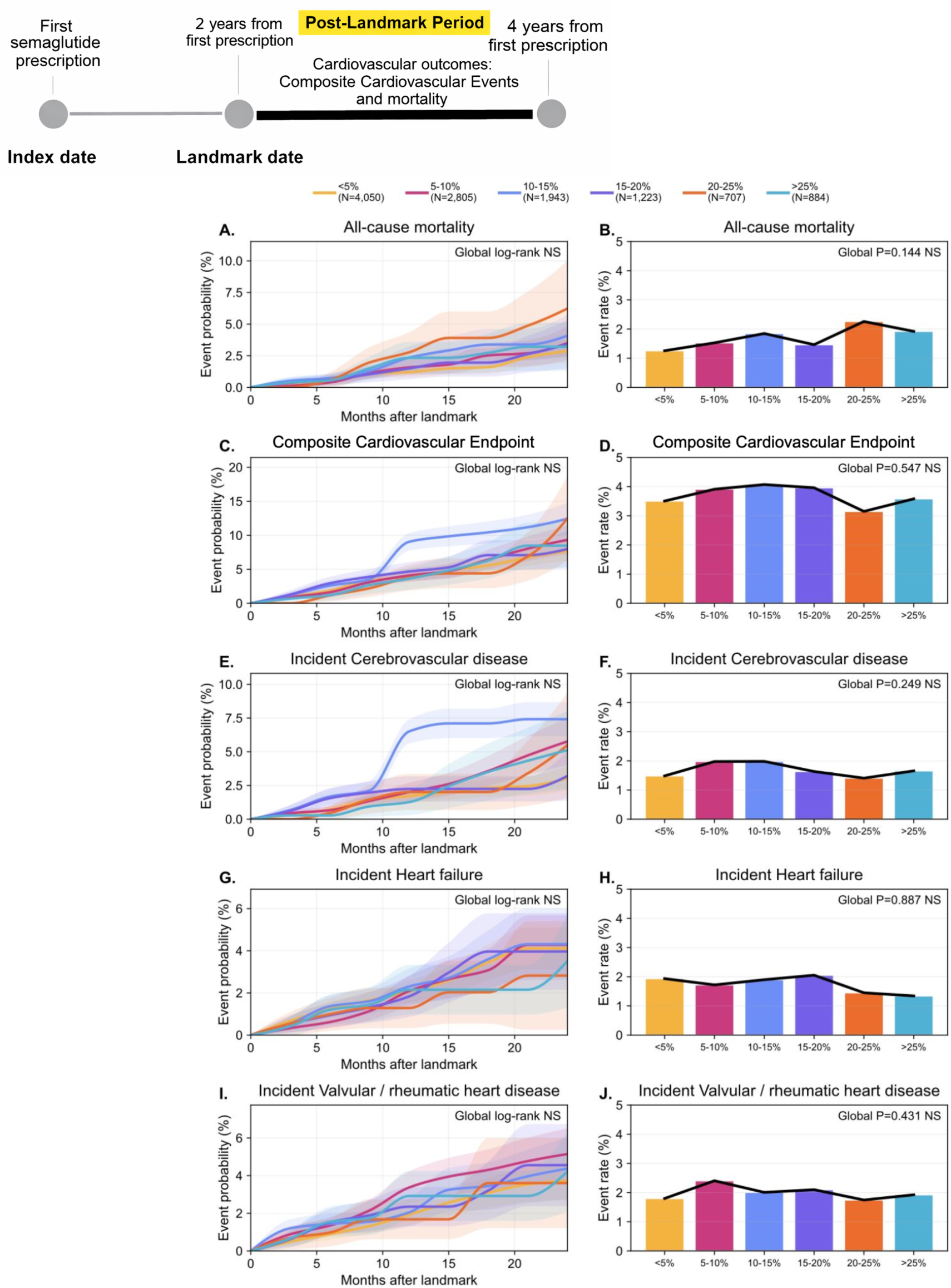
Semaglutide 2-year max weight-loss landmark analysis in cardiovascular conditions subgroup. Patients were categorized according to the maximum percentage reduction in body weight achieved before the 2-year landmark as <5% (*n* = 4,050), 5–10% (*n* = 2,805), 10–15% (*n* = 1,943), 15–20% (*n* = 1,223), 20–25% (*n* = 707), or >25% (*n* = 884), and cardiovascular outcomes were assessed only after the landmark among patients event-free through that timepoint. Left-column panels show cumulative post-landmark event probability over follow-up for all-cause mortality (A), composite cardiovascular events (C), incident cerebrovascular disease (E), incident heart failure (G), and incident valvular/rheumatic heart disease (I). Right-column panels show the corresponding post-landmark event rates across weight-loss categories for all-cause mortality (B), composite cardiovascular events (D), incident cerebrovascular disease (F), incident heart failure (H), and incident valvular/rheumatic heart disease (J), with global *P* values indicated. In contrast to the dose-defined analysis, achieved weight-loss strata did not show a consistent monotonic association with subsequent cardiovascular risk, and the representative global comparisons shown were not significant.

Part of the explanation may lie in the fact that weight loss was a moving summary of the first 2 years, whereas drug exposure fell sharply thereafter. When stratified by maximum dose achieved, semaglutide prescribing declined more sharply after the landmark among patients who had reached higher doses than among those who remained in the low-dose group, indicating that more intensive treatment during the first 2 years (landmark period) did not necessarily translate into sustained exposure in the following post-landmark period (**Fig. 6a**). A similar pattern was observed across weight-loss strata, with semaglutide prescriptions dropping markedly after the landmark (**Fig. 6b**). Median post-landmark prescription counts fell to 0 or 1 in every group, with the steepest declines in the higher-response strata: −4 [IQR −8 to −1] in the 15 to 20% group, −5 [IQR −9 to −1] in the 20 to 25% group, and −4 [IQR −8 to −1] in the >25% group (**Table S4**).

**Fig. 6.**
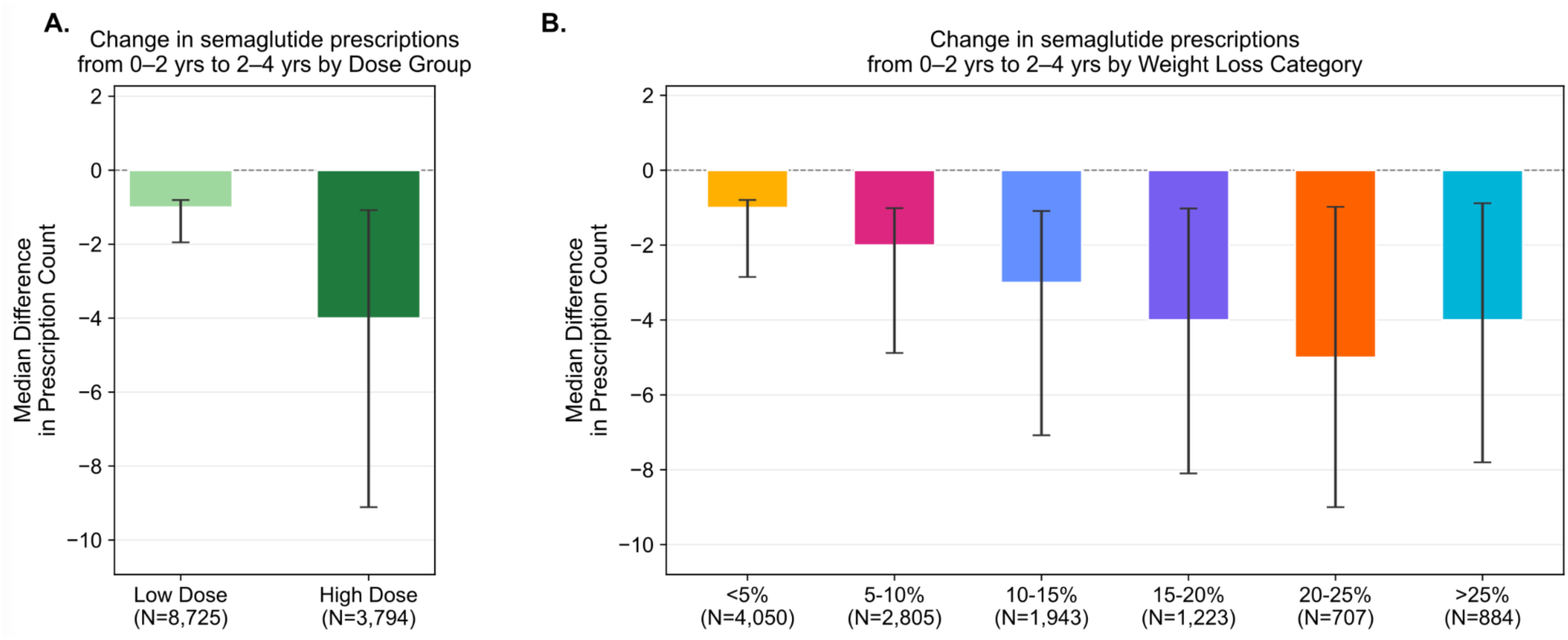
Reduction in semaglutide prescriptions in the two years following the initial treatment period, stratified by (A) maximum dose achieved and (B) maximum weight loss attained. Bars represent the median per-patient difference in prescription count between the 0–2 year and 2–4-year windows; error bars denote the interquartile range. Patients achieving higher doses or greater weight loss show larger declines in prescription fills, consistent with treatment tapering or discontinuation following peak therapeutic response.

Thus, maximum weight loss accrued during the landmark period did not correspond to sustained semaglutide prescriptions thereafter. Across all weight-loss strata, median prescription counts after maximum weight loss attainment were only 0 or 1, indicating that the subsequent outcome window was generally characterized by sparse semaglutide exposure (**Table S4**). Consistent with this context, a concurrent analysis anchored at the time of maximum weight loss showed that greater achieved weight loss was not associated with lower cardiovascular risk over the next 24 months (**Fig. 7**). Rather, all-cause mortality and the composite cardiovascular endpoint showed modest upward separation across increasing weight-loss categories, with the highest event probabilities generally observed among patients with the largest weight reductions (P > 0.05, **Fig. 7a-d**). Incident cerebrovascular disease followed a similar directional trend, whereas incident heart failure and incident valvular or rheumatic heart disease showed no consistent gradient across weight-loss strata (**Fig. 7e-j**). Together, these findings indicate that achieved weight loss alone did not recapitulate the favorable cardiovascular pattern observed across attained semaglutide dose tiers, particularly in a setting where semaglutide prescribing after maximum weight loss was minimal.

**Fig. 7.**
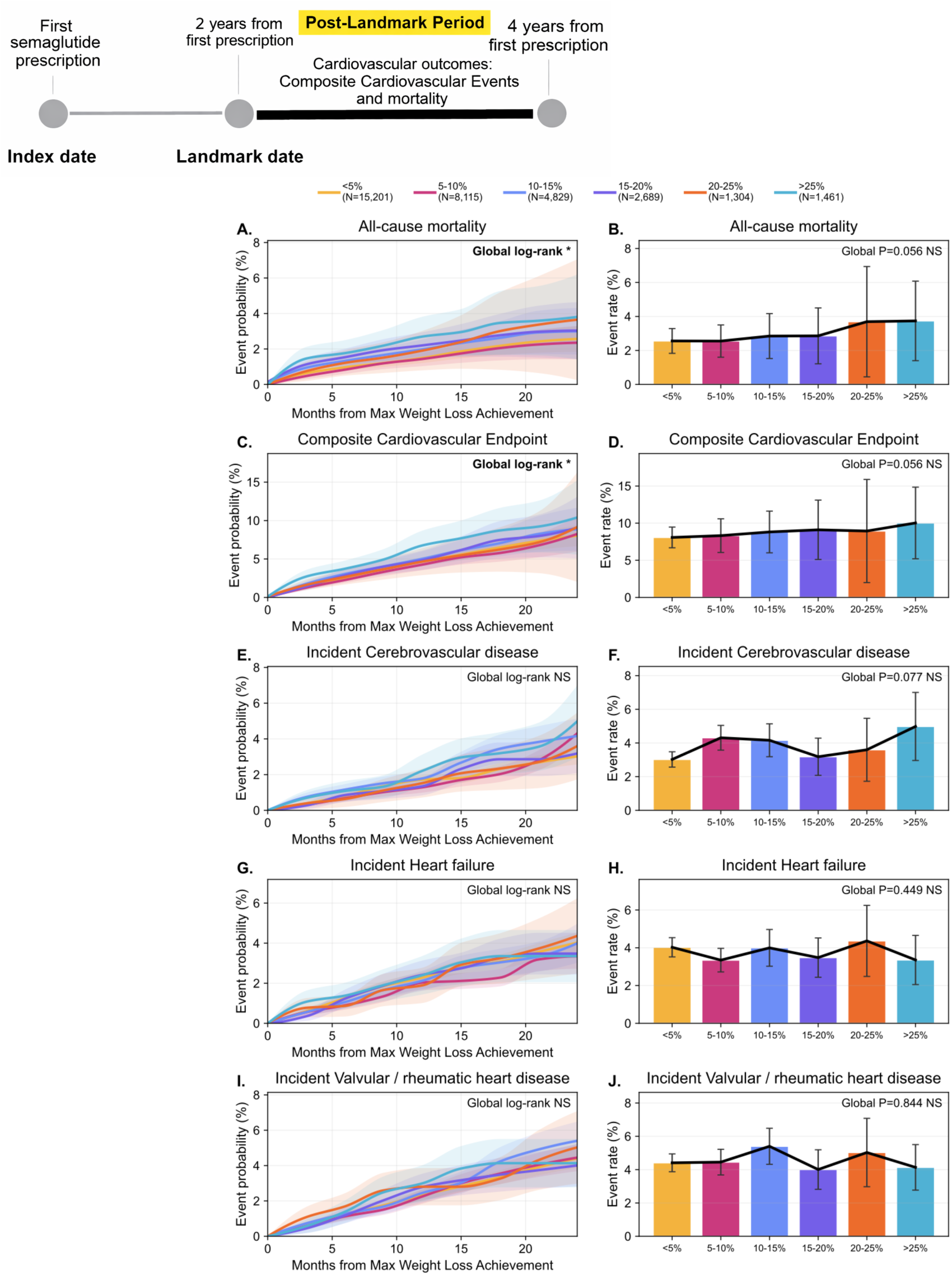
Concurrent relationship between achieved weight loss and cardiovascular outcomes during semaglutide treatment. Patients with at least one cardiovascular condition were categorized by the maximum percentage reduction in body weight achieved within a 2-year observation window as <5% (n=15,201), 5–10% (n=8,115), 10–15% (n=4,829), 15–20% (n=2,689), 20–25% (n=1,304), or >25% (n=1,461). For each patient, outcomes were tracked from the date they achieved their maximum weight loss, with follow-up extending up to 24 months thereafter. Left-column panels show cumulative event probability and right-column panels show the corresponding 24-month event rates for all-cause mortality (A, B), composite cardiovascular events (C, D), incident cerebrovascular disease (E, F), incident heart failure (G, H), and incident valvular/rheumatic heart disease (I, J). Error bars represent 95% confidence intervals derived from Greenwood’s formula; global log-rank P values are shown. In contrast to the dose-defined analysis, greater weight loss was associated with incrementally higher cardiovascular event rates across most outcomes — a pattern consistent with reverse causation, whereby patients with greater underlying cardiometabolic burden tend to achieve more pronounced weight loss on semaglutide during the concurrent observation window.

### Matched comparator analyses placed the semaglutide cardiovascular signal in the broader clinical context of antidiabetic medicines

Semaglutide carried a broader favorable cardiovascular signal in matched comparator analyses. In 1:1 propensity-score-matched comparisons against metformin, the pooled *all cardiovascular conditions* subgroup contained 47,199 patients per arm and was closely balanced for age at index (59.4 versus 59.9 years), female representation (62.8% versus 61.3%), baseline BMI (35.6 versus 35.3 kg/m^2^) and baseline type 2 diabetes prevalence (25.8% versus 26.0%) (**Table 2**). Across individual cardiovascular subgroups, mean age ranged from approximately 59 to 67 years, baseline BMI from 33.8 to 36.3 kg/m^2^, and baseline type 2 diabetes prevalence from 25.4% in aortic disease to 50.8% in heart failure, indicating broadly balanced matching across a clinically heterogeneous population.

**Table 2.** Baseline characteristics of propensity score–matched semaglutide and metformin cohorts across cardiovascular disease subgroups. For each baseline cardiovascular subgroup, patients initiating semaglutide were matched 1:1 to metformin-treated patients using propensity scores estimated from age, sex, baseline BMI and T2DM status. Matched cohort sizes, age at index date (mean ± SD), proportion female, baseline BMI (mean ± SD) and proportion with type 2 diabetes mellitus (T2DM) at baseline are shown.

| Baseline CV Group | Matched Number of unique patients | Semaglutide age at index, mean (SD) | Metformin age at index, mean (SD) | Semaglutide female, % | Metformin female, % | Semaglutide baseline BMI, mean (SD) | Metformin baseline BMI, mean (SD) | Semaglutide T2DM, % | Metformin T2DM, % |
| --- | --- | --- | --- | --- | --- | --- | --- | --- | --- |
| All CV conditions | 47199 | 59.4 (12.9) | 59.9 (13.3) | 62.8 | 61.3 | 35.6 (6.0) | 35.3 (6.0) | 25.8 | 26 |
| Aortic disease | 899 | 67.0 (10.8) | 67.5 (10.5) | 35.4 | 31.4 | 34.5 (5.5) | 34.3 (5.6) | 25.4 | 27.7 |
| Arrhythmia / Conduction disorders | 9590 | 63.6 (13.6) | 64.2 (14.7) | 56.9 | 54.3 | 35.1 (5.7) | 34.8 (6.0) | 29.8 | 30.4 |
| Cardiomyopathy | 1494 | 62.0 (13.0) | 62.4 (13.1) | 48.7 | 44 | 35.3 (5.8) | 34.7 (6.2) | 35.3 | 35.4 |
| Cerebrovascular disease | 2821 | 65.8 (11.8) | 66.1 (12.6) | 61.6 | 60.9 | 34.2 (5.7) | 34.2 (5.6) | 39.5 | 40.3 |
| Heart failure | 3777 | 65.9 (12.0) | 66.7 (12.6) | 53.2 | 48.7 | 36.2 (6.0) | 35.4 (6.6) | 50.8 | 50.2 |
| Hypertension | 40773 | 59.8 (12.6) | 60.4 (13.1) | 62 | 60.5 | 35.8 (5.9) | 35.5 (6.0) | 27.8 | 28.3 |
| Ischemic heart disease | 9203 | 66.4 (10.3) | 66.6 (10.5) | 44.8 | 41.8 | 34.6 (5.7) | 34.2 (5.7) | 38.7 | 37.8 |
| Peripheral / Vascular disease atherosclerosis | 3425 | 66.8 (11.3) | 67.2 (11.9) | 59.4 | 58.5 | 34.5 (6.1) | 33.8 (5.9) | 43.2 | 42.5 |
| Valvular / Rheumatic heart disease | 3082 | 65.2 (13.0) | 65.7 (13.8) | 60.4 | 58.4 | 34.5 (5.8) | 34.2 (6.0) | 34.7 | 35.1 |
| Venous thromboembolism | 2750 | 58.6 (13.6) | 59.4 (14.9) | 64.7 | 61.2 | 36.3 (6.1) | 35.9 (6.3) | 29.7 | 30.5 |

Analyzed from the time of treatment initiation (**Fig. 8a**), semaglutide was associated with lower approximate 2-year event risk than metformin for all-cause mortality (1.7% versus 3.4%; P < 0.001), the composite cardiovascular endpoint (9.7% versus 13.3%; P < 0.001), incident ischemic heart disease (8.0% versus 8.6%; P = 0.002), incident cerebrovascular disease (3.5% versus 4.8%; P < 0.001), incident hypertension (23.8% versus 40.5%; P < 0.001) and incident arrhythmias or conduction disorders (8.5% versus 10.4%; P < 0.001) (**Fig. 8b, Fig. S4a**). Broadly favorable patterns were also observed when comparing semaglutide to dipeptidyl peptidase-4 (DPP-4) inhibitors (**Fig. 8c, Fig. S4b**) or sodium-glucose cotransporter-2 (SGLT2) inhibitors (**Fig. 8d, Fig. S4c**). These comparator analyses support an overall favorable cardiovascular association for semaglutide.

**Fig. 8.**
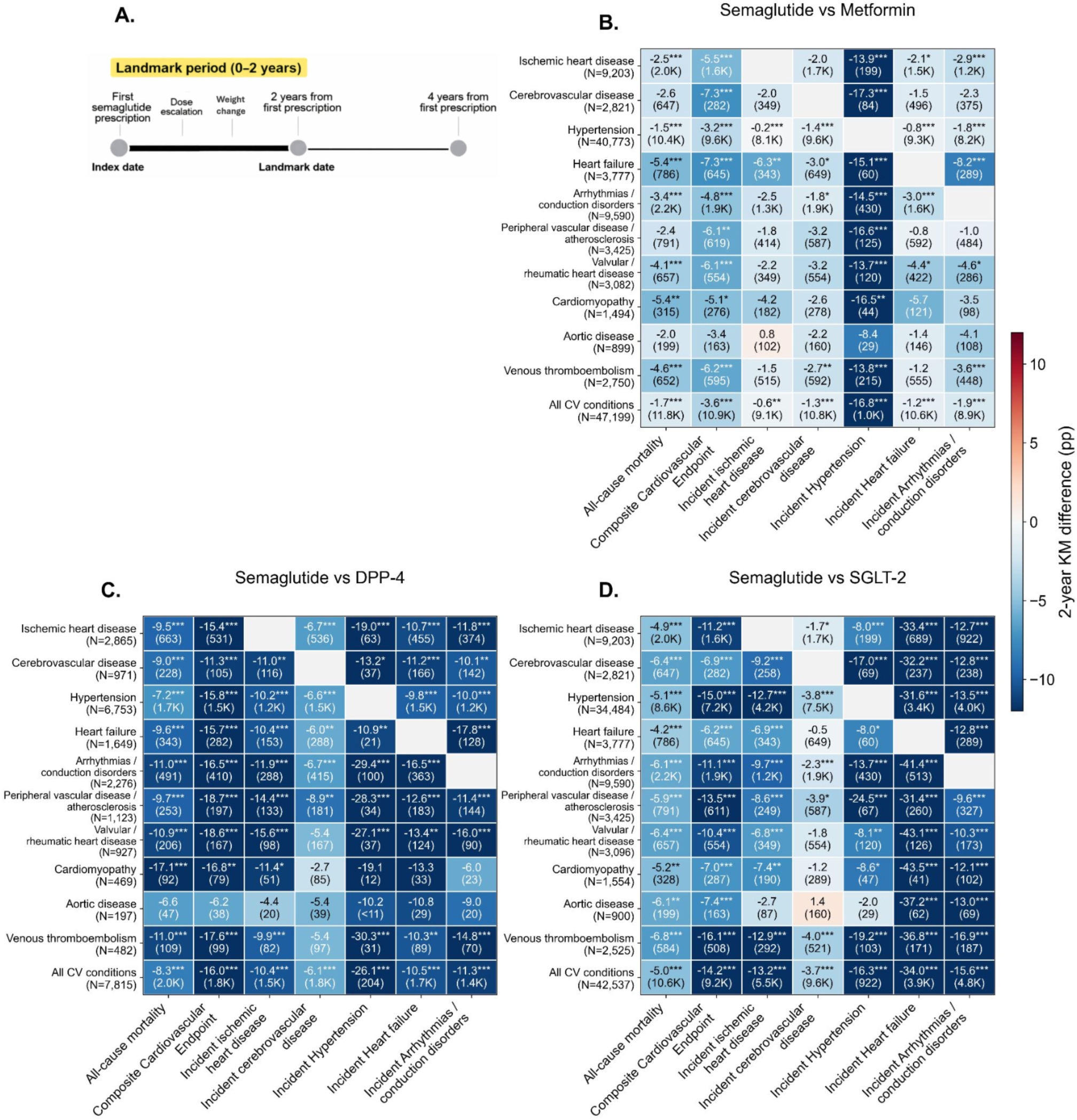
Semaglutide vs comparator anti-diabetic drugs’ absolute 2-year event risk differences across baseline cardiovascular burden subgroups. (A) Study design schematic highlighting the 0–2-year landmark period. (B) Semaglutide vs Metformin; (C) Semaglutide vs DPP-4 inhibitors; (D) Semaglutide vs SGLT-2 inhibitors. In B–D, heatmaps show absolute 2-year event risk differences across baseline cardiovascular burden subgroups. Rows indicate baseline cardiovascular burden subgroups, whereas columns indicate all-cause mortality, composite cardiovascular endpoint, and incident cardiovascular outcomes. Cell values represent the difference in approximate 2-year event risk between semaglutide and the respective comparator, expressed in percentage points (pp). Asterisks denote statistical significance based on log-rank tests using Fisher’s method (* p<0.05, ** p<0.01, *** p<0.001). Results for additional cardiovascular outcomes are shown in **Fig. S4**. **Table S6** lists the constituent medications within the DPP-4 inhibitors and SGLT2 inhibitors comparator groups. **Table S7** provides the full definitions of baseline cardiovascular burden groups. **Table S9** provides the component code lists used for the composite cardiovascular endpoint.

### Whole-body GLP1R geography provides biologic plausibility for organ-directed effects

The landmark analysis raised a biological question: if differences in subsequent cardiovascular outcomes among semaglutide-treated patients were not fully explained by the magnitude of weight loss achieved during the first 2 years, what aspects of GLP1R biology might account for this apparent organ-directed effect? We hypothesized that cell-type-specific receptor prevalence and downstream signaling readiness may be important determinants, and to test this idea, we generated a schematic model of *GLP1R engagement potential (GEP)* from a whole-body single-cell atlas (**Fig. 9a**). This model showed that the pancreas had the highest prevalence-weighted GEP (1.944%), but the heart emerged as the next most prominent tissue by GEP (0.729%) and, notably, as the largest aggregate GLP1R+ reservoir by *absolute target load (ATL*; 19,913), marginally exceeding the brain (19,217) and approximately doubling the pancreas (9,967) (**Fig. 9b**, **Table 3**). Thus, while the pancreas remained the dominant prevalence-weighted site of GLP1R signal, the heart stood out as a potential extra-pancreatic target organ.

**Fig. 9.**
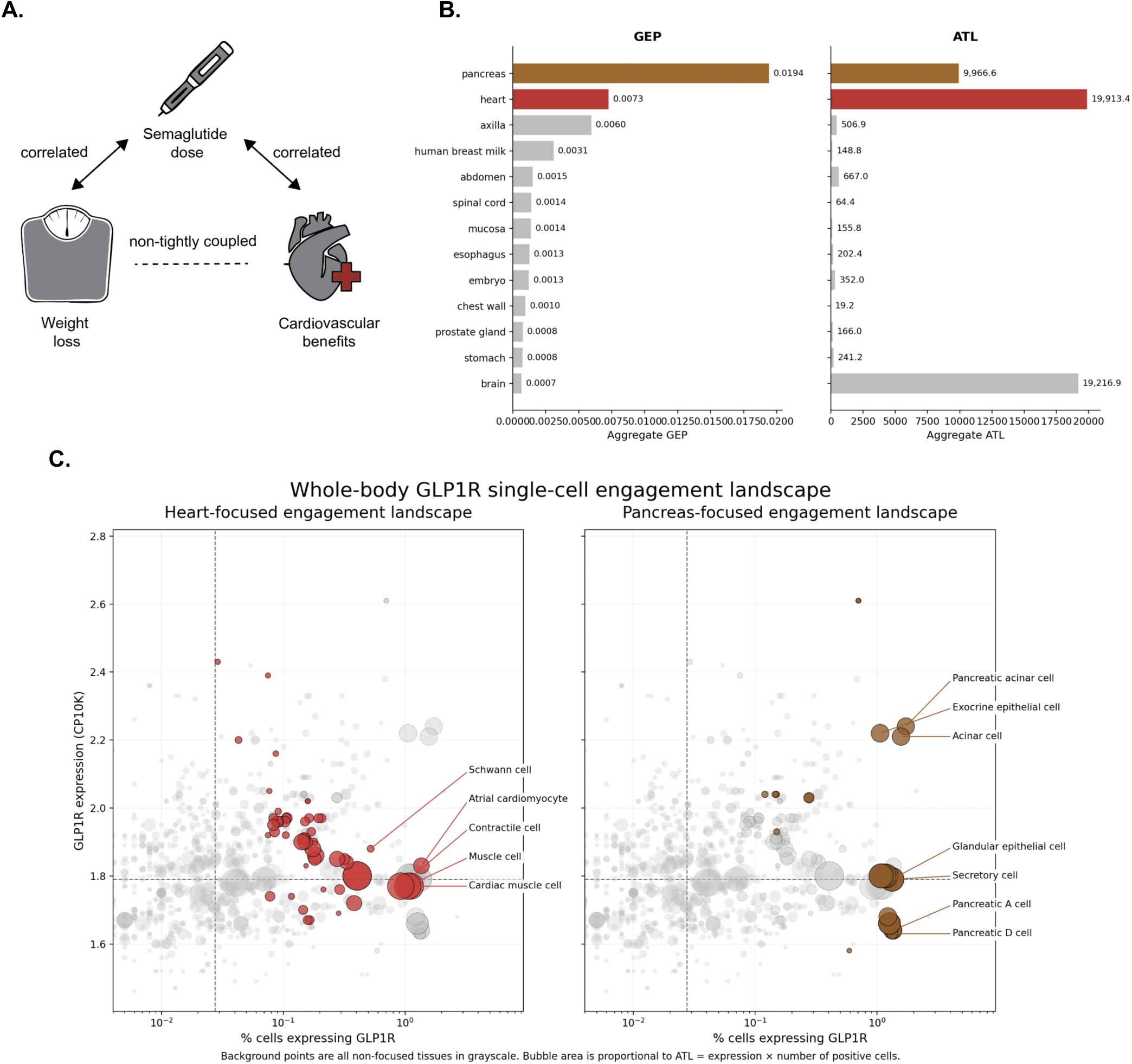
Whole-body GLP1R receptor geography distinguishes prevalence-weighted engagement from absolute target burden. **A.** Schematic overview of relationship between semaglutide dosage, weight loss and cardiovascular benefits **B.** *Semaglutide engagement across cardiac cell types stratified by GLP1R Engagement Potential (GEP; expression × positive-cell fraction).* Schematic illustrating a hypothesis-generating model in which increasing semaglutide exposure may broaden engagement from high-GEP cardiac cell types at lower doses to additional medium- and low-to-mid-GEP cell types at higher doses, contingent on the presence of downstream signaling and contextual gene programs. GEP is used as a prevalence-weighted proxy for receptor availability across heart cell types. This model is intended to motivate interrogation of dose-dependent cardiac target engagement and does not represent a validated pharmacokinetic–pharmacodynamic relationship. In addition to GEP which is the primary metric, the other metric computed is the Absolute Target Load (ATL; expression × number of GLP1R-positive cells). Tissues are ordered by descending GEP. Pancreas shows the highest prevalence-weighted GLP1R signal, whereas heart shows the largest transcript-weighted GLP1R-positive reservoir. **C,** Heart-focused single-cell engagement landscape. Each point represents a cell type, positioned by the percentage of cells expressing GLP1R on the x axis and GLP1R expression on the y axis; bubble area is proportional to ATL. Cardiomyocyte-related populations, including atrial cardiomyocytes, cardiac muscle cells, muscle cells and contractile cells, combine modest expression with appreciable prevalence, whereas Schwann cells show higher expression intensity but lower prevalence. Also shown alongside is the Pancreas-focused single-cell engagement landscape. Exocrine epithelial and acinar populations show both high prevalence and substantial expression, consistent with the dominance of pancreas in aggregate GEP. In b and c, heart or pancreas are highlighted, and all other tissues are shown in grayscale. Dashed lines indicate cohort-wide median values for percentage of cells expressing GLP1R and GLP1R expression.

**Table 3.** Single Cell RNA atlas of GLP1R Engagement Potential (GEP) and Absolute Target Load (ATL), with tissues sorted by decreasing GEP%.

| Tissue | Cell Count | Expression (CP10K) | % Expressing Cells | Positive Fraction | Cells Expr. GLP1R | GEP | GEP % | ATL |
| --- | --- | --- | --- | --- | --- | --- | --- | --- |
| pancreas | 512856 | 1.8 | 1.08 | 0.0108 | 5537 | 0.01944 | 1.944 | 9966.6 |
| heart | 2730947 | 1.8 | 0.405 | 0.00405 | 11063 | 0.00729 | 0.729 | 19913.4 |
| axilla | 84584 | 1.73 | 0.346 | 0.00346 | 293 | 0.0059858 | 0.59858 | 506.89 |
| human breast milk | 47514 | 1.71 | 0.183 | 0.00183 | 87 | 0.0031293 | 0.31293 | 148.77 |
| abdomen | 429748 | 1.68 | 0.092 | 0.00092 | 397 | 0.0015456 | 0.15456 | 666.96 |
| spinal cord | 44639 | 1.79 | 0.081 | 0.00081 | 36 | 0.0014499 | 0.14499 | 64.44 |
| mucosa | 109366 | 2.05 | 0.069 | 0.00069 | 76 | 0.0014145 | 0.14145 | 155.8 |
| esophagus | 155431 | 2.2 | 0.059 | 0.00059 | 92 | 0.001298 | 0.1298 | 202.4 |
| embryo | 277234 | 2 | 0.063 | 0.00063 | 176 | 0.00126 | 0.126 | 352 |
| chest wall | 19459 | 1.75 | 0.057 | 0.00057 | 11 | 0.0009975 | 0.09975 | 19.25 |
| prostate gland | 207838 | 2.05 | 0.039 | 0.00039 | 81 | 0.0007995 | 0.07995 | 166.05 |
| stomach | 303597 | 1.93 | 0.041 | 0.00041 | 125 | 0.0007913 | 0.07913 | 241.25 |
| brain | 26689529 | 1.78 | 0.04 | 0.0004 | 10796 | 0.000712 | 0.0712 | 19216.88 |
| adrenal gland | 244608 | 1.73 | 0.04 | 0.0004 | 98 | 0.000692 | 0.0692 | 169.54 |
| adipose tissue | 448846 | 1.87 | 0.037 | 0.00037 | 164 | 0.0006919 | 0.06919 | 306.68 |
| lymph node | 1731461 | 1.73 | 0.033 | 0.00033 | 576 | 0.0005709 | 0.05709 | 996.48 |
| musculature | 508632 | 1.98 | 0.028 | 0.00028 | 144 | 0.0005544 | 0.05544 | 285.12 |
| kidney | 1245614 | 1.85 | 0.028 | 0.00028 | 352 | 0.000518 | 0.0518 | 651.2 |
| exocrine gland | 193695 | 2.02 | 0.022 | 0.00022 | 42 | 0.0004444 | 0.04444 | 84.84 |
| liver | 1444734 | 1.73 | 0.024 | 0.00024 | 346 | 0.0004152 | 0.04152 | 598.58 |
| breast | 3100226 | 1.84 | 0.022 | 0.00022 | 696 | 0.0004048 | 0.04048 | 1280.64 |
| intestine | 322530 | 1.85 | 0.02 | 0.0002 | 64 | 0.00037 | 0.037 | 118.4 |
| lung | 4526576 | 1.83 | 0.02 | 0.0002 | 909 | 0.000366 | 0.0366 | 1663.47 |
| eye | 7700797 | 1.75 | 0.02 | 0.0002 | 1548 | 0.00035 | 0.035 | 2709 |
| tendon of semitendinosus | 10533 | 1.77 | 0.019 | 0.00019 | 2 | 0.0003363 | 0.03363 | 3.54 |
| neck | 19097 | 1.65 | 0.016 | 0.00016 | 3 | 0.000264 | 0.0264 | 4.95 |
| colon | 812980 | 1.83 | 0.012 | 0.00012 | 99 | 0.0002196 | 0.02196 | 181.17 |
| cortex | 149400 | 1.65 | 0.013 | 0.00013 | 19 | 0.0002145 | 0.02145 | 31.35 |
| chest | 15413 | 1.55 | 0.013 | 0.00013 | 2 | 0.0002015 | 0.02015 | 3.1 |
| tongue | 38749 | 1.47 | 0.013 | 0.00013 | 5 | 0.0001911 | 0.01911 | 7.35 |
| testis | 18528 | 1.58 | 0.011 | 0.00011 | 2 | 0.0001738 | 0.01738 | 3.16 |
| small intestine | 1191162 | 1.71 | 0.01 | 0.0001 | 119 | 0.000171 | 0.0171 | 203.49 |
| cerebrospinal fluid | 72565 | 1.76 | 0.008 | 8.00E-05 | 6 | 0.0001408 | 0.01408 | 10.56 |
| spleen | 537601 | 1.69 | 0.008 | 8.00E-05 | 42 | 0.0001352 | 0.01352 | 70.98 |
| skin of body | 817861 | 1.91 | 0.007 | 7.00E-05 | 57 | 0.0001337 | 0.01337 | 108.87 |
| esophagogastric junction | 11883 | 1.54 | 0.008 | 8.00E-05 | 1 | 0.0001232 | 0.01232 | 1.54 |
| endocrine gland | 654345 | 1.7 | 0.006 | 6.00E-05 | 38 | 0.000102 | 0.0102 | 64.6 |
| blood | 25136830 | 1.67 | 0.005 | 5.00E-05 | 1330 | 8.35E-05 | 0.00835 | 2221.1 |
| ureter | 41757 | 1.56 | 0.005 | 5.00E-05 | 2 | 7.80E-05 | 0.0078 | 3.12 |
| ovary | 346569 | 1.73 | 0.004 | 4.00E-05 | 14 | 6.92E-05 | 0.00692 | 24.22 |
| bone marrow | 727397 | 1.65 | 0.004 | 4.00E-05 | 30 | 6.60E-05 | 0.0066 | 49.5 |
| urinary bladder | 36100 | 1.51 | 0.003 | 3.00E-05 | 1 | 4.53E-05 | 0.00453 | 1.51 |
| fallopian tube | 238661 | 1.98 | 0.002 | 2.00E-05 | 4 | 3.96E-05 | 0.00396 | 7.92 |
| large intestine | 422380 | 1.77 | 0.002 | 2.00E-05 | 8 | 3.54E-05 | 0.00354 | 14.16 |
| pleural fluid | 83186 | 1.46 | 0.002 | 2.00E-05 | 2 | 2.92E-05 | 0.00292 | 2.92 |
| uterus | 456208 | 2.05 | 0.001 | 1.00E-05 | 4 | 2.05E-05 | 0.00205 | 8.2 |
| vasculature | 349181 | 1.69 | 0.001 | 1.00E-05 | 4 | 1.69E-05 | 0.00169 | 6.76 |
| placenta | 561073 | 1.68 | 0.001 | 1.00E-05 | 7 | 1.68E-05 | 0.00168 | 11.76 |
| omentum | 213994 | 1.62 | 0.001 | 1.00E-05 | 2 | 1.62E-05 | 0.00162 | 3.24 |
| bladder organ | 68317 | 1.46 | 0.001 | 1.00E-05 | 1 | 1.46E-05 | 0.00146 | 1.46 |
| hindlimb | 85565 | 1.46 | 0.001 | 1.00E-05 | 1 | 1.46E-05 | 0.00146 | 1.46 |

Within the heart, the most prevalent GLP1R+ populations were cardiomyocytes, particularly atrial cardiomyocytes, followed by endocardial cells, pericytes, and cardiac blood vessel endothelial cells (**Fig. 9c; Table S5**). Schwann cells and specific cardiac immune-cell populations showed some of the highest per-cell expressions but were rare and ultra-rare, respectively. The GLP1R+ rare cardio-immune cells included CD4-positive alpha-beta T cells and mature natural killer T cells. Among cardiac myocytes, regular atrial cardiac myocytes showed GLP1R expression of 1.83 CP10K, with 992 expressing cells and 1.364% of cells positive, yielding a salience score of 2.492, whereas regular ventricular cardiac myocytes showed comparable per-cell expression intensity (1.72 CP10K) but substantially lower prevalence, with 926 expressing cells and only 0.382% of cells positive, yielding a lower salience score of 0.659. A second ventricular population, ventricular cardiac muscle cells, showed slightly higher per-cell expression (2.03 CP10K) but extremely sparse positivity (0.116%; salience 0.236). The dominant contribution in the pancreas arose from the exocrine compartment including acinar cells, which had the top-ranked GEP status across all analyzed organs of the human body (**Fig. 9d**).

Consistent with these insights from single cell analysis, whole-body bulk RNA sequencing data shows that GLP1R expression was highest in the pancreas, followed next by the heart, and then by selected neuroendocrine tissues (**Fig. S5**). Specifically, heart muscle ranked among the highest non-pancreatic tissues for GLP1R expression, significantly exceeding most peripheral organs, including lung, liver, kidney, and adipose tissue. Within cardiac compartments, GLP1R expression showed meaningful signal in atrial tissue and, to a lesser extent, ventricular tissue, with distributions indicating heterogeneous but reproducible expression across samples. Atrial samples (n=432) and left ventricular samples (n=432, 550) showed median TPM was 1.01 in atrial tissues (Q1–Q3, 0.56–1.47) versus 0.44–0.46 in left ventricular tissues (Q1–Q3, 0.17–0.87). Triangulating bulk RNA and single-cell RNA-seq, atrial tissue showed a stronger GLP1R signal than ventricular tissue, with the bulk distributions shifted upward in atrium across the full range of samples: median TPM was 1.01 in atrial tissues (Q1–Q3, 0.56–1.47) versus 0.44–0.46 in left ventricular tissues (Q1–Q3, 0.17–0.87). Together, these data indicate that the stronger atrial GLP1R signal in bulk tissue is better explained by higher prevalence of GLP1R-positive cardiomyocytes in the atrium than by markedly higher per-cell expression intensity.

Collectively, these data establish that while the pancreas dominates GLP1R expression, the heart represents a prominent extra-pancreatic site of GLP1R transcription, supporting the plausibility of cardiac cells as biologically relevant targets for GLP1 receptor agonists which may contribute to their broader effects beyond body-weight reduction.

## Discussion

### Semaglutide appears to write on more than one physiologic axis

The most important lesson of these data is that semaglutide seems to leave at least two signatures on human physiology. One signature is immediately visible: higher attained dose tracked closely with greater weight loss across the first 2 years after initiation. The second is quieter, but potentially more consequential: later cardiovascular benefit aligned more closely with dose attained than with the magnitude of weight loss itself. If weight loss were the dominant mediator of cardiovascular protection in this setting, one would expect a graded decline in post-landmark events across progressively deeper weight-loss strata. We did not observe that pattern. Instead, conventional metabolic markers such as HbA1c and blood pressure improved monotonically with weight loss, whereas post-landmark cardiovascular outcomes did not. In this cohort, body weight change therefore behaved as an important pharmacodynamic readout, but not as a complete surrogate for the biology most relevant to later cardiovascular risk.

The prescribing trajectory after the landmark sharpens that distinction. Weight loss was a cumulative summary of the first 2 years, whereas semaglutide prescribing attenuated substantially in the subsequent interval in which cardiovascular outcomes were assessed. In that setting, a patient’s maximum weight loss may memorialize earlier exposure without faithfully representing the therapeutic state that followed. That temporal mismatch provides one explanation for why weight-loss magnitude tracked systemic metabolic improvement yet failed to organize later cardiovascular risk as cleanly as dose attainment did.

### Cardiac GLP1R geography offers biologic plausibility for organ-directed effects

Our atlas-based analyses offer a biologically plausible framework for this dissociation. The pancreas remained the dominant prevalence-weighted site of GLP1R signal, as expected for an incretin therapy. But the heart emerged immediately behind it by GLP1R engagement potential and, notably, as the largest transcript-weighted reservoir of GLP1R-positive cells (**Fig. 9**, **Table 3**). Within that reservoir, signal was not confined to a single rare niche. It extended across cardiomyocyte and contractile populations, endothelial and endocardial compartments, stromal and perivascular states, and selected immune populations. These data do not demonstrate direct cardiac causality. They do, however, make it biologically plausible that the cardiovascular effects of semaglutide are written partly through organ-directed biology in the heart and vasculature, and not solely through the arithmetic of kilograms lost.

That broader view is also consistent with the emerging clinical literature. Semaglutide has shown cardiovascular and heart-failure benefits across multiple contexts, including obesity with established cardiovascular disease, obesity-related HFpEF, chronic kidney disease with diabetes, oral semaglutide cardiovascular outcomes, and symptomatic peripheral artery disease^3–10^. Our data suggest that these benefits may be better understood as the net result of several partially overlapping programs, including systemic metabolic change, vascular-interface effects, and myocardial or immunometabolic signaling. An extended mechanistic interpretation of the whole-body atlas, cardiac cell-state programs, and functional-readiness modules is provided in the Supplementary Discussion (**Figs. S5**-**S6**). Finally, differential expression of GLP1R in the heart also motivates future analyses of cardiac remodeling in semaglutide-treated patients through radiological diagnostic imaging data collected before and after semaglutide treatment initiation.

### Implications for endpoint selection, dose optimization and trial design

The matched comparator analyses place this within-drug observation into a broader therapeutic frame. From treatment initiation, semaglutide showed a favorable cardiovascular association relative to metformin, DPP-4 inhibitors, and SGLT2 inhibitors across several major endpoints. Taken together with the landmark analyses, the implication is not that weight loss is irrelevant. Rather, it is that cardiovascular medicine should resist reducing the value of semaglutide to a single visible phenotype. Weight loss may be the most conspicuous manifestation of drug activity, but it is not necessarily the most discriminating measure of organ benefit.

This distinction matters for both clinical practice and drug development. In routine care, incretin therapies are often titrated and compared through the lens of body-weight reduction. Yet the outcomes that matter most to patients with cardiovascular disease are myocardial infarction, stroke, heart failure, disability, and death. If these data are confirmed prospectively, then dose attainment, exposure patterns and organ-specific endpoints should stand alongside weight loss, rather than behind it, in the optimization of semaglutide and in the design of next-generation trials. The broader opportunity is methodological as well: a whole-body spatial intelligence framework that brings together federated longitudinal EHR data, therapeutic exposure, physiologic biomarkers, and molecular atlases may help reveal therapeutic value axes that conventional obesity endpoints do not fully capture.

### Limitations and future directions

This study has important limitations. As an observational analysis of de-identified EHR data, it remains vulnerable to residual confounding, treatment-selection bias, unequal measurement density, unmeasured adherence, and misclassification of outcomes defined from diagnosis codes rather than adjudicated events. Dose was inferred from prescriptions rather than verified administration. The marked attenuation of prescribing after the landmark further complicates interpretation of exposure during the later risk window. The transcriptomic analyses provide biologic plausibility, not mechanistic proof: single-cell and bulk RNA measurements do not establish receptor occupancy, downstream pathway activation, or causal mediation in human cardiac tissue. The metrics GEP and ATL should be interpreted as atlas-derived pharmacologic heuristics rather than literal measures of in vivo receptor occupancy or absolute organ-level target burden. In particular, ATL is influenced by uneven cell recovery and differential susceptibility of cell types to dissociation, capture and sequencing in single-cell RNA-sequencing datasets, and therefore should not be treated as a direct estimate of absolute cellular proportions across tissues. Accordingly, these metrics are most useful for prioritizing candidate tissues and cell compartments for semaglutide responsiveness, rather than for making exact quantitative claims about organ-level pharmacologic exposure.

The absence of a monotonic cardiovascular gradient across weight-loss strata should therefore not be read as evidence that large semaglutide-mediated weight loss is harmful. It should be read more carefully, and more usefully, as evidence that weight-loss magnitude alone does not fully explain the later cardiovascular signal observed here. Even with those caveats, a coherent picture emerges. Semaglutide appears to act not only as an agent of body-mass reduction, but as a therapy whose clinical signature extends into cardiovascular biology. One part of that signature is visible on the scale. Another may be written more quietly in the heart, the vessel wall, and the clinical arc of risk over time. The next generation of cardiometabolic trials will need to read both.

## Methods

### Data Source and Study Design

We conducted a retrospective cohort study using deidentified longitudinal electronic health record data from the nSights Federated EHR Network^11^. All analyses were conducted on deidentified data.

### Study Cohorts and Exposure Definitions

From approximately 29 million patients, 505,874 had at least one semaglutide prescription record, of whom 269,390 initiated semaglutide between March 2018 and January 2024; 47,199 of these had baseline cardiovascular disease (see below). Medication exposure was ascertained from prescription or medication event records using the recorded event timestamp. The index date was defined as the date of the first qualifying prescription for the cohort-defining medication or medication class. Follow-up data were available through January 2026. To create incident-user cohorts with minimal treatment contamination, patients were excluded if they had exposure to metformin, DPP-4 inhibitors, or SGLT2 inhibitors during the 365 days before the index date or at any time after the index date. Constituent medications within DPP-4 and SGLT2 groups are listed in **Table S6**. Other GLP-1RA medications (tirzepatide, liraglutide, dulaglutide, exenatide, albiglutide, lixisenatide) were used as exclusion exposures when defining the semaglutide cohort.

### Demographic and Clinical Covariates

Demographic data included age, sex, and death date. Age at index was obtained from the index medication record. Baseline BMI was defined as the BMI measurement closest to the index date, recorded from 365 days before through 14 days after the index date. Baseline weight for the weight-loss analyses was defined separately as the weight measurement closest to the index date, recorded from 90 days before through 14 days after the index date.

Baseline type 2 diabetes was defined by the presence of at least three encounters carrying a type 2 diabetes code on three distinct dates at any time before the index date. The same logic was used to define prespecified baseline cardiovascular comorbidity groups. These groups were ischemic heart disease, cerebrovascular disease, hypertension, heart failure, arrhythmias or conduction disorders, peripheral vascular disease, or atherosclerosis, valvular or rheumatic heart disease, cardiomyopathy, aortic disease, and venous thromboembolism. Diagnosis definitions were based on prespecified ICD-9 and ICD-10 code prefixes; the full diagnosis code sets are provided in **Table S7**. An all-cardiovascular conditions subgroup, defined as the union of these groups, served as the primary analytic cohort for the dose- and weight-loss-based landmark analyses; individual cardiovascular subgroups were examined separately in the comparative outcome analyses. We also calculated the interval between the index date and the most recent qualifying preindex diagnosis code for each cardiovascular subgroup; these summaries are provided in **Table S8**.

### Cardiovascular Outcomes

The primary comparative outcomes were all-cause mortality, composite cardiovascular events (all-cause death, MI, or stroke), and incident cardiovascular diagnoses. All-cause mortality was defined by the recorded death date. The composite cardiovascular endpoint was defined as all-cause death or the first post-index diagnosis consistent with acute myocardial infarction, acute coronary syndrome, or cerebrovascular event; component ICD-9 and ICD-10 code definitions are provided in **Table S9**. Incident cardiovascular outcomes included incident ischemic heart disease, cerebrovascular disease, hypertension, heart failure, arrhythmias or conduction disorders, peripheral vascular disease, or atherosclerosis, valvular or rheumatic heart disease, cardiomyopathy, aortic disease, and venous thromboembolism. For each incident outcome analysis, patients with evidence of that same condition before the start of follow-up were excluded.

### Dose–Weight-Loss Relationship

To characterize the dose–response relationship between semaglutide and weight reduction, we examined the distribution of maximum percent body-weight change achieved within the 2-year landmark window according to the maximum semaglutide dose reached during that period. Dose categories corresponded to labeled dosing increments for injectable semaglutide formulations (Ozempic and Wegovy): 0.25 mg, 0.5 mg, 1.0 mg, 1.7 mg, 2.0 mg, and 2.4 mg or greater. Observed semaglutide dose values were used when directly available in the medication record, and missing dose values were extracted from medication descriptions using a large language model-assisted dose-mapping workflow based on GPT-OSS-20B^15^. The resulting normalized dose values were used to assign patients to labeled dosing categories. This workflow increased dose ascertainment coverage from 56.0% to 98.2% of semaglutide prescription records. Manual review of 148 unique medication descriptions confirmed correct dose assignment in every case; the only apparent ambiguities (13 descriptions, 8.8%) arose from multi-dose pen devices listing two possible doses (e.g., “0.25 mg or 0.5 mg”), which were assigned the higher of the two labeled doses as a convention. A linear regression was fitted to the mean maximum weight-loss percentage at each dose level to quantify the overall dose-response trend.

### Landmark Analyses of Cardiovascular Outcomes by Dose and Weight Loss at 2 Years

We performed within-drug landmark analyses to evaluate the association of dose intensity and weight loss with subsequent cardiovascular outcomes among semaglutide-treated patients. The landmark was defined at 2 years after the index date. Outcomes were assessed only after the landmark date, and patients who experienced the outcome of interest on or before the landmark were excluded from the corresponding analysis. In the post-landmark period, patients were followed until the outcome of interest, death, or the last observed clinical record, whichever occurred first.

For dose-based analyses, we used all prescriptions from the index date through the landmark to define the maximum dose reached by the landmark. Semaglutide dose was dichotomized as low (0.25–1.0 mg) or high (1.7 mg or greater). Patients were considered analyzable if follow-up extended through the landmark and at least one dose-bearing prescription was available by that time point.

For weight-loss-based analyses, baseline weight was defined as the measurement closest to the index date, from 90 days before through 14 days after index. Post-index weight change was evaluated beginning 15 days after index through the landmark to avoid overlap between baseline ascertainment and follow-up weight assessment. Maximum percent body-weight change was calculated as the difference between baseline weight and the lowest observed weight before the landmark, divided by baseline weight, multiplied by 100. Patients were categorized into prespecified strata: less than 5%, 5–10%, 10–15%, 15–20%, 20–25%, and 25% or greater. Analyses were restricted to patients with an available baseline weight and at least one follow-up weight before the landmark.

For both dose and weight-loss landmark analyses, cardiovascular outcomes were evaluated using cumulative event curves over the 24-month post-landmark period and overall post-landmark event proportions.

### Post-Landmark Metabolic Surrogates, Prescription Continuity, and Sensitivity Analyses

Post-landmark metabolic surrogates included HbA1c and systolic and diastolic blood pressure. For each patient, the post-landmark value was defined as the closest measurement to the landmark date within the 24-month post-landmark period. Change from baseline was calculated as the difference between the post-landmark value and the most recent pre-treatment measurement. These surrogates were summarized across weight-loss strata.

To assess semaglutide prescription continuity relative to the landmark, we computed per-patient prescription counts in two windows: the pre-landmark window spanning from the first semaglutide prescription date to the 2-year landmark date, and the post-landmark window spanning the 24 months following the landmark date (truncated at the last observation date). The per-patient difference in prescription count (post minus pre) was computed and summarized as median [IQR] stratified by dose group and weight-loss category.

In addition to the shared 2-year landmark, two sensitivity analyses were performed: the first anchored follow-up to the date of first attainment of the patient’s maximum semaglutide dose, and the second to the date of maximum body-weight reduction. In both cases, outcomes were evaluated over the subsequent 24 months to test whether the patterns observed with the fixed landmark were robust when aligned to the time points at which peak exposure or peak response was achieved.

### Propensity-Matched Comparative Cardiovascular Analyses

For comparator analyses, three additional mutually exclusive incident-user cohorts were constructed for metformin, DPP-4 inhibitors, and SGLT2 inhibitors using the same index-date and exclusion logic described above. Within each baseline cardiovascular subgroup, we performed pairwise comparisons anchored on semaglutide as the primary exposure cohorts. Separate analyses were conducted for semaglutide versus metformin, DPP-4 inhibitors, and SGLT2 inhibitors. Propensity scores were estimated with logistic regression using age at index, sex, baseline BMI, and baseline type 2 diabetes status. Patients were matched 1:1 without replacement using nearest-neighbor matching and a caliper of 0.2 on the propensity score scale.

Follow-up for comparative outcome analyses began on the index date. Patients were followed until the outcome of interest, death, or the last observed clinical record. For incident cardiovascular outcomes, patients with prevalent disease of the same type at baseline were excluded from that specific analysis. We summarized matched cohort size, event counts, person-time, and cumulative 2-year event estimates using Kaplan-Meier methods and log-rank P values.

### Analysis of GLP1R Expression Using Single-Cell and Bulk Tissue Atlases

Single-cell expression of *GLP1R* was obtained from the CELLxGENE data portal^16^ (https://cellxgene.cziscience.com/), leveraging 1556 publicly available human single-cell and single-nucleus RNA sequencing datasets corresponding to 86 million cells from 11,633 human donors. Using the CELLxGENE Census, GLP1R expression was obtained across available cell types. Gene expression values were extracted as normalized counts (counts per 10,000 [CP10K], log-transformed where applicable) and analyzed at the cell-type level using curated annotations provided within each dataset. For each cell type, *GLP1R* expression was summarized as both mean normalized expression and the proportion of expressing cells (non-zero counts). Where multiple datasets contributed to the same tissue, results were aggregated to ensure robustness across cohorts, and analyses were restricted to cell types with adequate representation to minimize sparsity-driven artifacts.

Bulk tissue-level expression of GLP1R was obtained from the Human Protein Atlas (proteinatlas.org). Normalized transcript expression values (e.g., nTPM) were retrieved across human tissues, with particular emphasis on cardiac subregions where available.

### Single Cell RNA-sequencing profiling of GLP1R across human body tissues

Single-cell GLP1R atlas data were analyzed as follows. For each tissue-cell annotation, Expression (CP10K), number of GLP1R-positive cells, and percentage of expressing cells were analyzed.

The primary metric was *GLP1R Engagement Potential (GEP)*, defined as Expression (CP10K) × (% expressing cells / 100). Conceptually, GEP is a prevalence-weighted expression metric: it becomes high when a cell class has both appreciable GLP1R transcript abundance and a meaningful proportion of cells that are GLP1R-positive. It is therefore useful for ranking which cellular compartments are most likely to be engaged under a broad systemic exposure model because it balances intensity and prevalence rather than privileging either one alone. A compartment with very high expression in only a tiny number of cells may not rank as highly by GEP as a compartment with slightly lower expression but a much broader GLP1R-positive fraction.

The other salient metric was *Absolute Target Load (ATL)*, defined as the product of per-cell GLP1R expression and the absolute number of GLP1R-positive cells in that compartment. This is computed as Expression (CP10K) × NumberPositiveCells, where NumberPositiveCells is the number of cells expressing GLP1R in that tissue-cell class. Conceptually, ATL estimates the total transcript-weighted burden of GLP1R-positive cells and is therefore a whole-compartment burden metric rather than a prevalence-weighted one. It becomes especially informative at the tissue or organ level, where a compartment may have only modest prevalence but still contain a very large absolute reservoir of GLP1R-positive cells because the total sampled population is large. For this reason, ATL is particularly useful for understanding overall organ-scale target burden and for distinguishing tissues that may represent major aggregate pharmacologic reservoirs even if their percentage of positive cells is not the highest.

Stable cell classes were filtered for visualization using Cell Count >= 100 and NumberPositiveCells >= 2.

### Statistical Analysis

Continuous variables were summarized as means with standard deviations or medians with interquartile ranges, as appropriate; categorical variables were summarized as counts and percentages. In the comparative cardiovascular outcome analyses, between-cohort differences were assessed using propensity score–matched Kaplan-Meier estimates of 2-year event risk; statistical significance in KM-difference heatmaps was determined by log-rank test. In the dose landmark analyses, cumulative event distributions were compared between high-dose and low-dose groups using log-rank tests, and post-landmark event rates were compared using relative risks with associated Wald p-values. In the weight-loss landmark analyses, cumulative event distributions and post-landmark event rates across weight-loss strata were compared using a global log-rank test. Trends in post-landmark metabolic surrogates (HbA1c, systolic and diastolic blood pressure) across weight-loss strata were assessed using one-way ANOVA. Analyses were performed in Python 3.13.1 using pandas 3.0.0, NumPy 2.4.2, SciPy 1.17.0, and Matplotlib 3.10.8; propensity score modeling, survival analyses and single-cell expression profiling were implemented within the same analytic pipeline.

## Data Source

This study analyzed de-identified EHR data from academic medical centers in the United States via the nference nSights Analytics Platform. Prior to analysis, all data underwent expert determination de-identification satisfying HIPAA Privacy Rule requirements (45 CFR §164.514(b)(1)), employing a multi-layered transformation approach for both structured data (cryptographic hashing of identifiers, date-shifting, geographic truncation) and unstructured clinical text (ensemble deep learning and rule-based methods with >99% recall for personally identifiable information detection)^17,18^. nference established secure data environments within each participating center, housing these de-identified patient data governed by expert determination. These de-identified data environments were specifically designed to enable data access and analysis without requiring Institutional Review Board oversight, approval, or exemption confirmation. Accordingly, informed consent and IRB review were not required for this study.

## Data Availability

This study involves the analysis of de-identified Electronic Health Record (EHR) data via the nference nSights Federated Clinical Analytics Platform (nSights). Data shown and reported in this manuscript were extracted from this environment using an established protocol for data extraction, aimed at preserving patient privacy. The data has been de-identified pursuant to an expert determination in accordance with the HIPAA Privacy Rule. Any data beyond what is reported in the manuscript, including but not limited to the raw EHR data, cannot be shared or released due to the parameters of the expert determination to maintain the data de-identification. The corresponding author should be contacted for additional details regarding nSights.

## De-identification and HIPAA compliance certification

Prior to analysis, all EHR data were de-identified under an expert determination consistent with the Health Insurance Portability and Accountability Act (HIPAA) Privacy Rule (45 CFR §164.514(b)(1)). The de-identification methodology employed a multi-layered transformation approach to both structured and unstructured data fields^17,18^. In structured data, direct identifiers including patient names and precise geographic locations were excluded entirely, while indirect identifiers underwent specific transformations: patient identifiers, medical record numbers, and accession numbers were replaced with one-way cryptographic hashes using confidential salts to preserve linkage across patient encounters; all dates were shifted backward by patient-specific random offsets (1–31 days) to preserve temporal relationships while obscuring exact event timing; the ZIP codes were truncated to two-digit state-level resolution; and continuous variables including age, height, weight, and body mass index were thresholded to prevent identification of extreme values (for example, ages ≥89 years transformed to ‘89+’ and BMI >40 transformed to ‘40+’). In unstructured clinical text, an ensemble de-identification system that combines attention-based deep learning models with rule-based methods achieved an estimated >99% recall for personally identifiable information (PII) detection, with detected identifiers replaced by plausible fictional surrogates^17^.

## Data Harmonization

To address heterogeneity in EHR data, we harmonized clinical variables including medications, anthropometric measurements, and diagnoses to standardized concepts. For medications, we first constructed a standardized drug concept database combining the nSights knowledge graph with RXNorm (https://www.nlm.nih.gov/research/umls/rxnorm/index.html) hierarchies to capture ingredient, brand, and dose-specific information^11^. EHR medication records were matched using a hierarchical approach prioritizing RXNorm codes when available, followed by ingredient-level matching, and finally natural language processing and pattern matching on free-text medication orders when structured codes were absent. For anthropometric measurements (height, weight, BMI), we created a unified vocabulary from SNOMED (https://www.snomed.org/, https://athena.ohdsi.org) and LOINC (https://loinc.org/) terminologies and matched EHR measurement descriptions using standardized text matching algorithms with abbreviation expansion and synonym resolution; ambiguous mappings were resolved using OpenAI GPT-4o (https://platform.openai.com/docs/models/gpt-4o) with summary statistics as context, followed by manual verification. For diagnoses, we developed a hierarchical disease concept database from the nSights knowledge graph and matched EHR diagnosis descriptions and codes by identifying the most specific common child concept in the hierarchy. This approach enabled consistent identification of clinical entities while preserving granularity where available.

## Acknowledgements

We thank the nference engineering team for the development of the nSights federated AI platform, and Patrick Lenehan for critical review and manuscript feedback.

## Supplementary Information

**Fig. S1.**
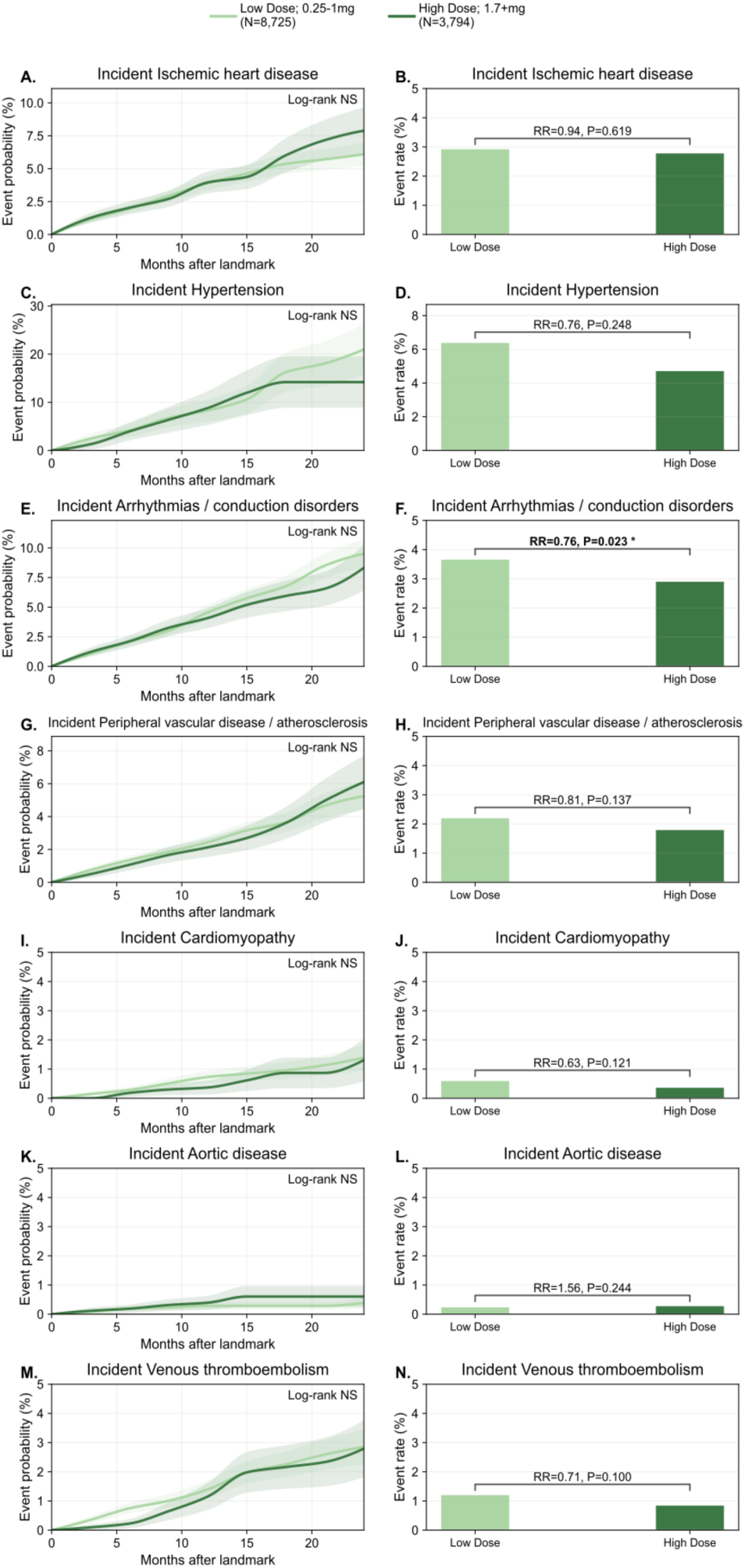
Higher maximum semaglutide dose was associated with lower post-landmark incidence of select cardiovascular conditions. Patients were grouped by the maximum semaglutide dose reached by the 2-year landmark as low dose (0.25–1.0 mg; n = 8,725) or high dose (≥1.7 mg; n = 3,794), and incident cardiovascular outcomes were assessed only after the landmark among patients event-free through that timepoint. Left-column panels show cumulative post-landmark event probability over follow-up for incident ischemic heart disease (A), incident hypertension (C), incident arrhythmias/conduction disorders (E), incident peripheral vascular disease/atherosclerosis (G), incident cardiomyopathy (I), incident aortic disease (K), and incident venous thromboembolism (M). Right-column panels show the corresponding post-landmark event rates with relative risks (RR) and nominal P values for incident ischemic heart disease (B), incident hypertension (D), incident arrhythmias/conduction disorders (F), incident peripheral vascular disease/atherosclerosis (H), incident cardiomyopathy (J), incident aortic disease (L), and incident venous thromboembolism (N).

**Fig. S2.**
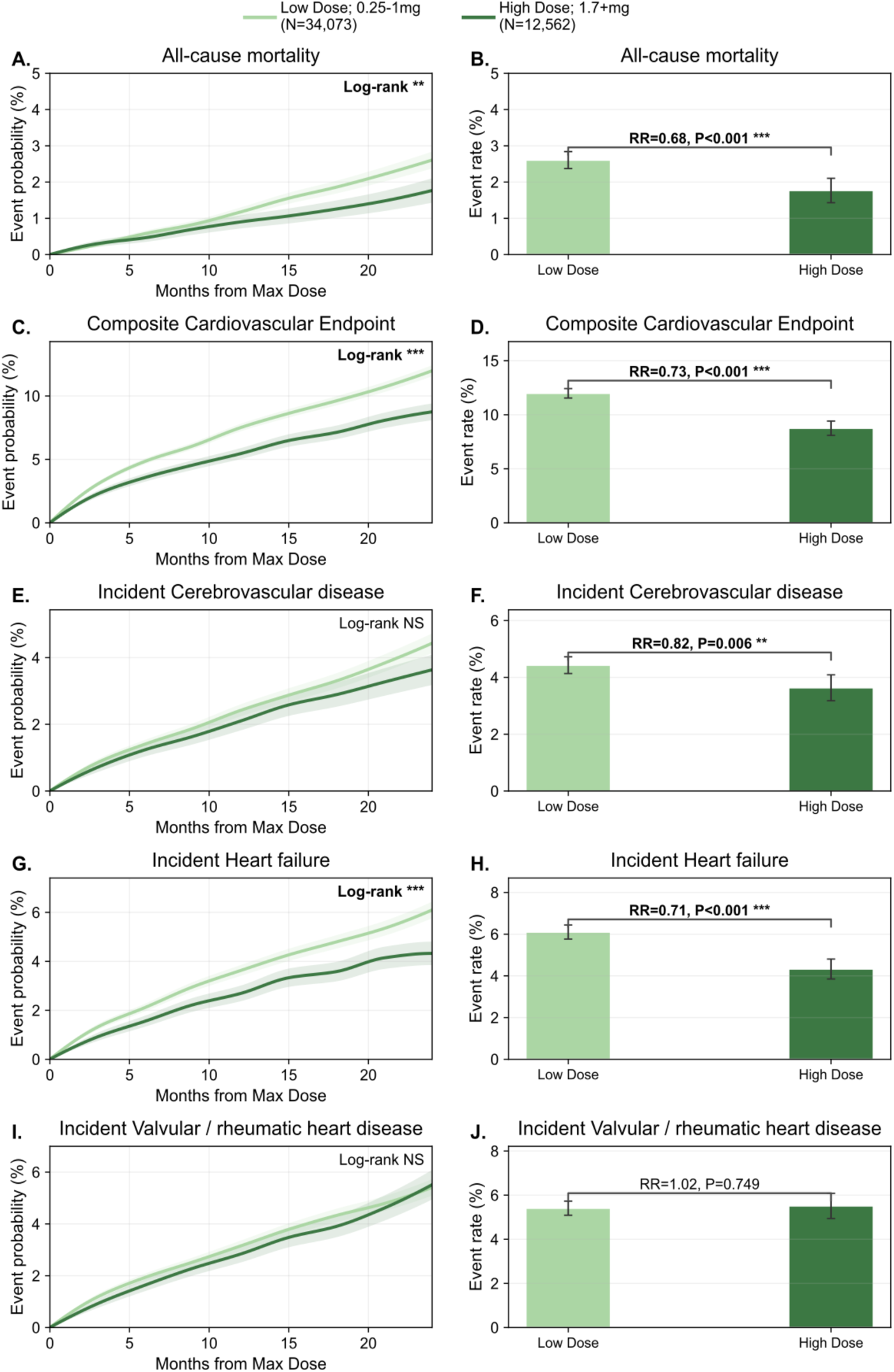
Higher maximum semaglutide dose was associated with lower cardiovascular risk. Patients with at least one cardiovascular condition were grouped by the maximum semaglutide dose reached within a 2-year observation window as low dose (0.25–1.0 mg; n=34,073) or high dose (≥1.7 mg; n=12,562). For each patient, outcomes were tracked from the date they first achieved their maximum dose, with follow-up extending up to 24 months thereafter. Left-column panels show cumulative event probability and right-column panels show the corresponding 24-month event rates for all-cause mortality (A, B), composite cardiovascular events (C, D), incident cerebrovascular disease (E, F), incident heart failure (G, H), and incident valvular/rheumatic heart disease (I, J). Error bars represent 95% confidence intervals derived from Greenwood’s formula; relative risks and nominal P values comparing high versus low dose are shown on the bar plots. Higher-dose semaglutide was associated with lower risk across all outcomes examined.

**Fig. S3.**
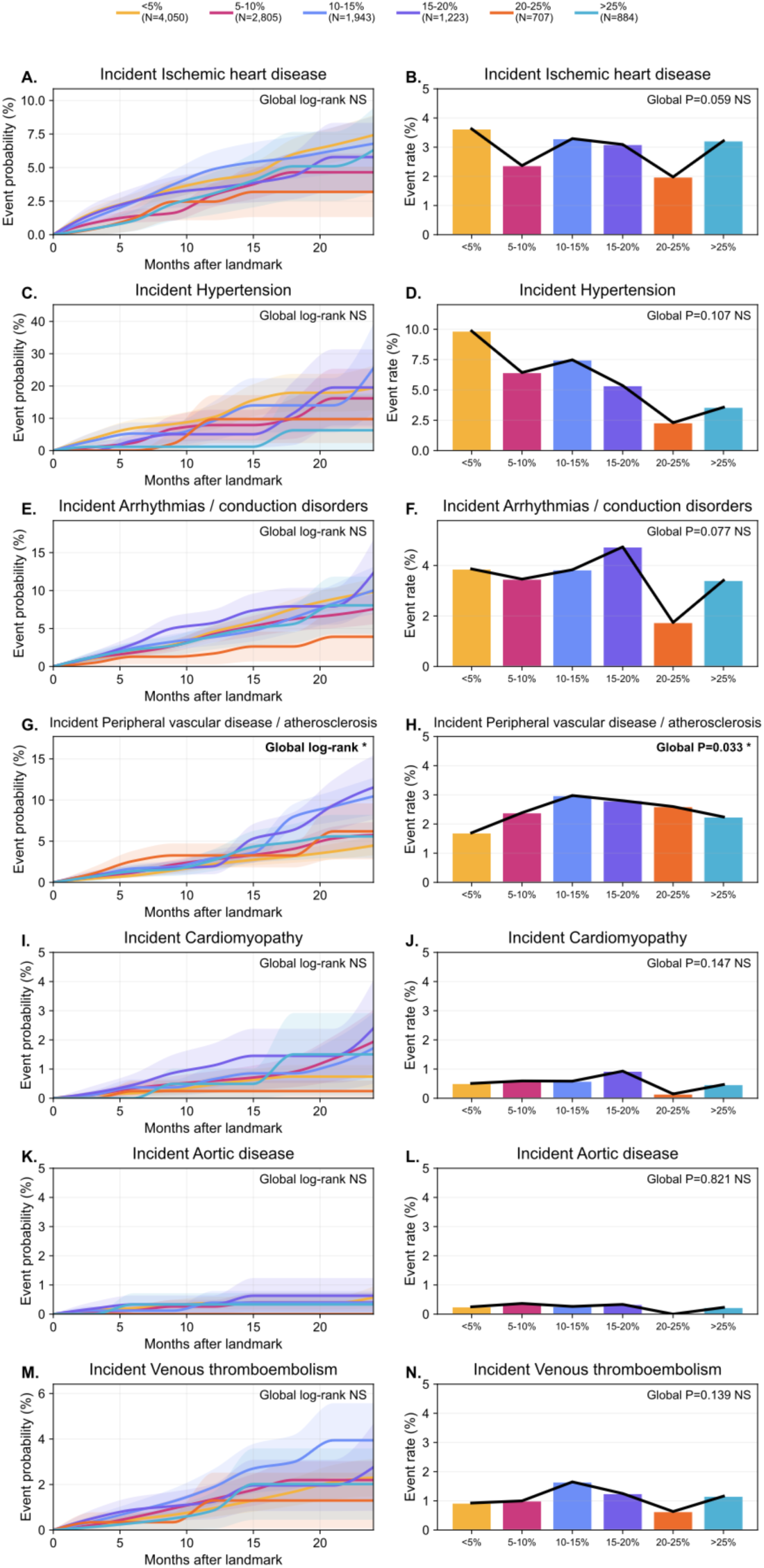
Semaglutide 2-year maximum weight-loss landmark analysis: incident cardiovascular conditions by weight-loss category. Patients were categorized according to the maximum percentage reduction in body weight achieved before the 2-year landmark as <5% (n = 4,050), 5–10% (n = 2,805), 10–15% (n = 1,943), 15–20% (n = 1,223), 20–25% (n = 707), or >25% (n = 884), and incident cardiovascular outcomes were assessed only after the landmark among patients event-free through that timepoint. Left-column panels show cumulative post-landmark event probability over follow-up for incident ischemic heart disease (A), incident hypertension (C), incident arrhythmias/conduction disorders (E), incident peripheral vascular disease/atherosclerosis (G), incident cardiomyopathy (I), incident aortic disease (K), and incident venous thromboembolism (M). Right-column panels show the corresponding post-landmark event rates across weight-loss categories for incident ischemic heart disease (B), incident hypertension (D), incident arrhythmias/conduction disorders (F), incident peripheral vascular disease/atherosclerosis (H), incident cardiomyopathy (J), incident aortic disease (L), and incident venous thromboembolism (N), with global P values indicated.

**Fig. S4.**
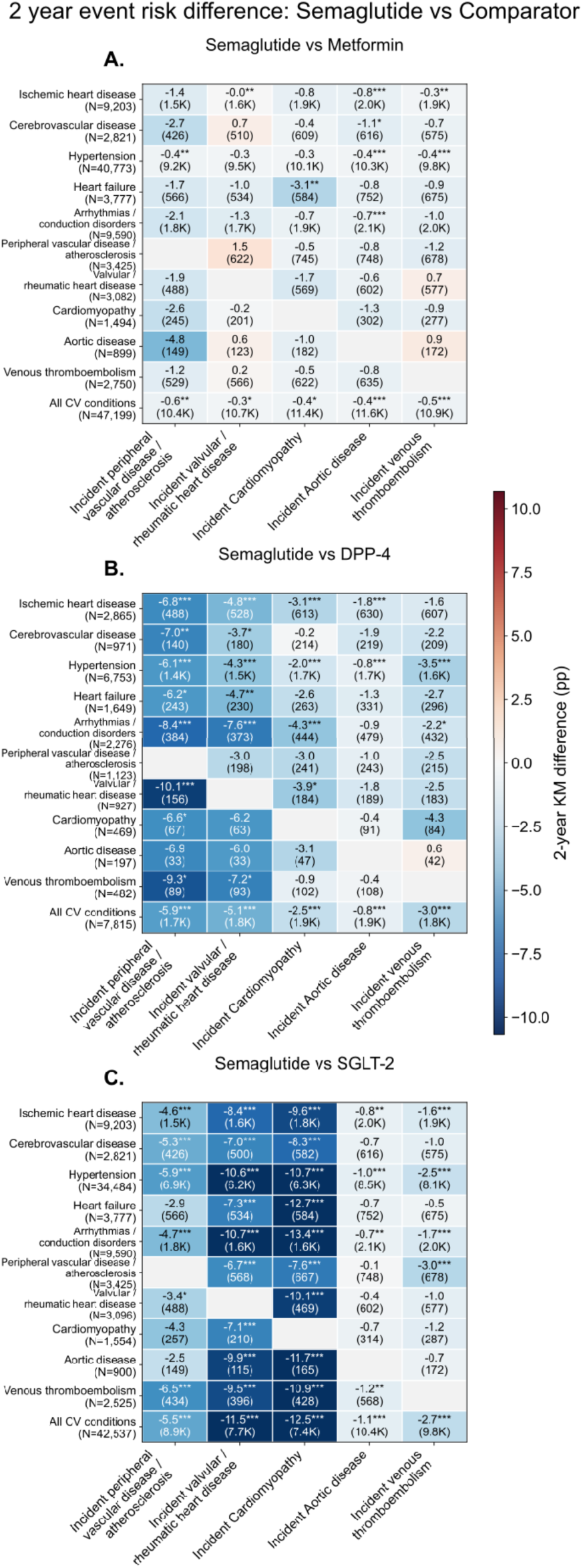
Semaglutide vs comparator anti-diabetic drugs’ absolute 2-year event risk differences across baseline cardiovascular burden subgroups for additional cardiovascular outcomes. (A) Semaglutide vs Metformin; (B) Semaglutide vs DPP-4 inhibitors; (C) Semaglutide vs SGLT-2 inhibitors. Heatmaps show absolute 2-year event risk differences across baseline cardiovascular burden subgroups. Rows indicate baseline cardiovascular burden subgroups, whereas columns indicate incident peripheral vascular disease/atherosclerosis, valvular/rheumatic heart disease, cardiomyopathy, aortic disease, and venous thromboembolism. Cell values represent the difference in approximate 2-year event risk between semaglutide and the respective comparator, expressed in percentage points (pp). Asterisks denote statistical significance (* p<0.05, ** p<0.01, *** p<0.001).

**Fig. S5.**
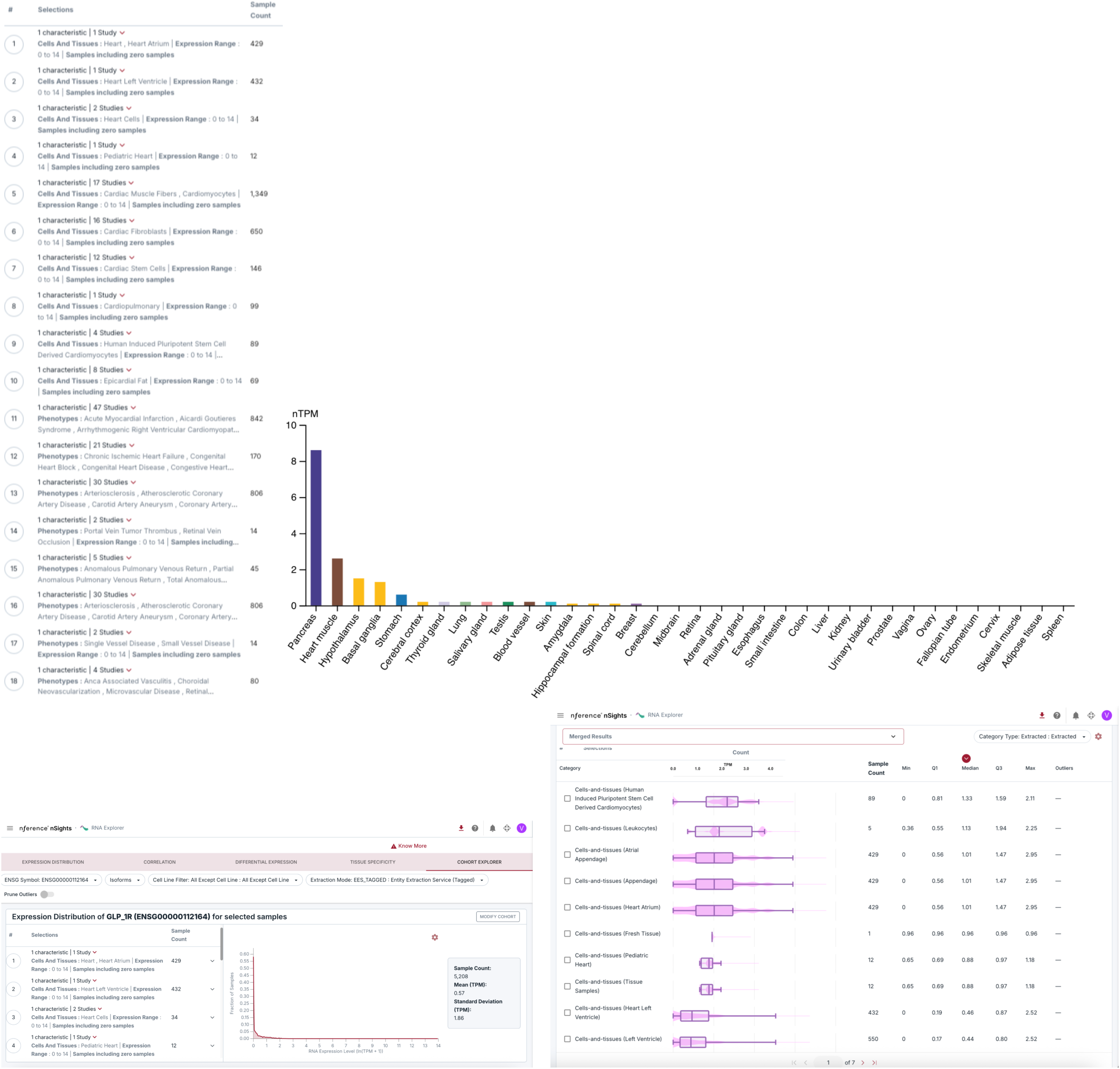
Bulk RNA expression profiling of GLP1R with Heart and Cardiovascular Samples in this study. https://nfer-workspaces.com/rna_lab/results?token=glp+1r&ranges=%5B0%2C0.013%2C1.004%2C6.651%5D&cell_line_data_source=not_cell_line&single_cell_data_source=not_single_cell&diff_data_type=gtex_tcga&input_type=gene&collections=%5Broot-cells-and-tissues%2Croot-diseases%5D&enriched_study=high&include_cohen_d_pos=true&include_cohen_d_neg=true&single_cell_details_sort=adj_score&tissue_spec_dataset=gtex&include_cohen_d_pos_tiss_spec=true&include_cohen_d_neg_tiss_spec=true&cluster_name=all&cluster_category=all&study_exp_range=%5B0%2C14%5D&includeSamples=all&ensg_symbol=ENSG00000112164&enst_symbol=none&extraction_mode=ees_tagged&corr_data_type=whole_group&data_type=cohort_info&groupA=_%24eyIxIjp7InNhbXBsZUNvdW50Ijo2Mzk5MCwiZXhwUmFuZ2UiOlswLDE0XSwiY2F0ZWdvcnlKb2luIjp7IndpdGhpbl9jYXRlZ29yeV9qb2luIjp7Im5mZXItc3R1ZGllcyI6InVuaW9uIiwibmZlci1waGVub3R5cGVzIjoidW5pb24iLCJuZmVyLWNlbGxzLWFuZC10aXNzdWVzIjoiaW50ZXJzZWN0aW9uIiwibmZlci1kcnVncyI6ImludGVyc2VjdGlvbiIsIm5mZXItZXRobmljaXR5IjoiaW50ZXJzZWN0aW9uIiwibmZlci1hZ2UiOiJpbnRlcnNlY3Rpb24iLCJuZmVyLWdlbmRlciI6ImludGVyc2VjdGlvbiIsInJhdy1tZXRhZGF0YSI6ImludGVyc2VjdGlvbiJ9fSwiYWxsU2FtcGxlcyI6ImFsbCIsImZpbHRlciI6eyJuZmVyLWdlbmRlciI6WyJtYWxlIl19LCJwb3NpdGlvbiI6MSwibXV0YXRpb25zIjpbXSwicmF3Q2hhcmFjdGVyaXN0aWNzIjp7fSwiYWxsU2VsZWN0ZWQiOmZhbHNlLCJzdHVkeUNvdW50IjoxNDc5LCJjaGFyYWN0ZXJpc3RpY3NDb3VudCI6MSwidHlwZSI6ImNvaG9ydCIsInRpdGxlIjoiMTogMSBjaGFyYWN0ZXJpc3RpYyB8IDE0NzkgU3R1ZGllcyJ9fQ%3D%3D&prune_outliers=false https://nfer-workspaces.com/rna_lab/results?token=glp+1r&ensg_symbol=ENSG00000112164&enst_symbol=none&ranges=%5B0%2C0.013%2C1.004%2C6.651%5D&default_bins=_%24eyJyYW5nZV9hX21pbiI6MCwicmFuZ2VfYV9tYXgiOjAuMDEzLCJyYW5nZV9iX21pbiI6MS4wMDQsInJhbmdlX2JfbWF4Ijo2LjY1MX0%3D&data_type=cohort_info&prune_outliers=false&groupA=69c2950750dd1bf75b8347ac&isMinifyGroupA=true

**Fig. S6.**
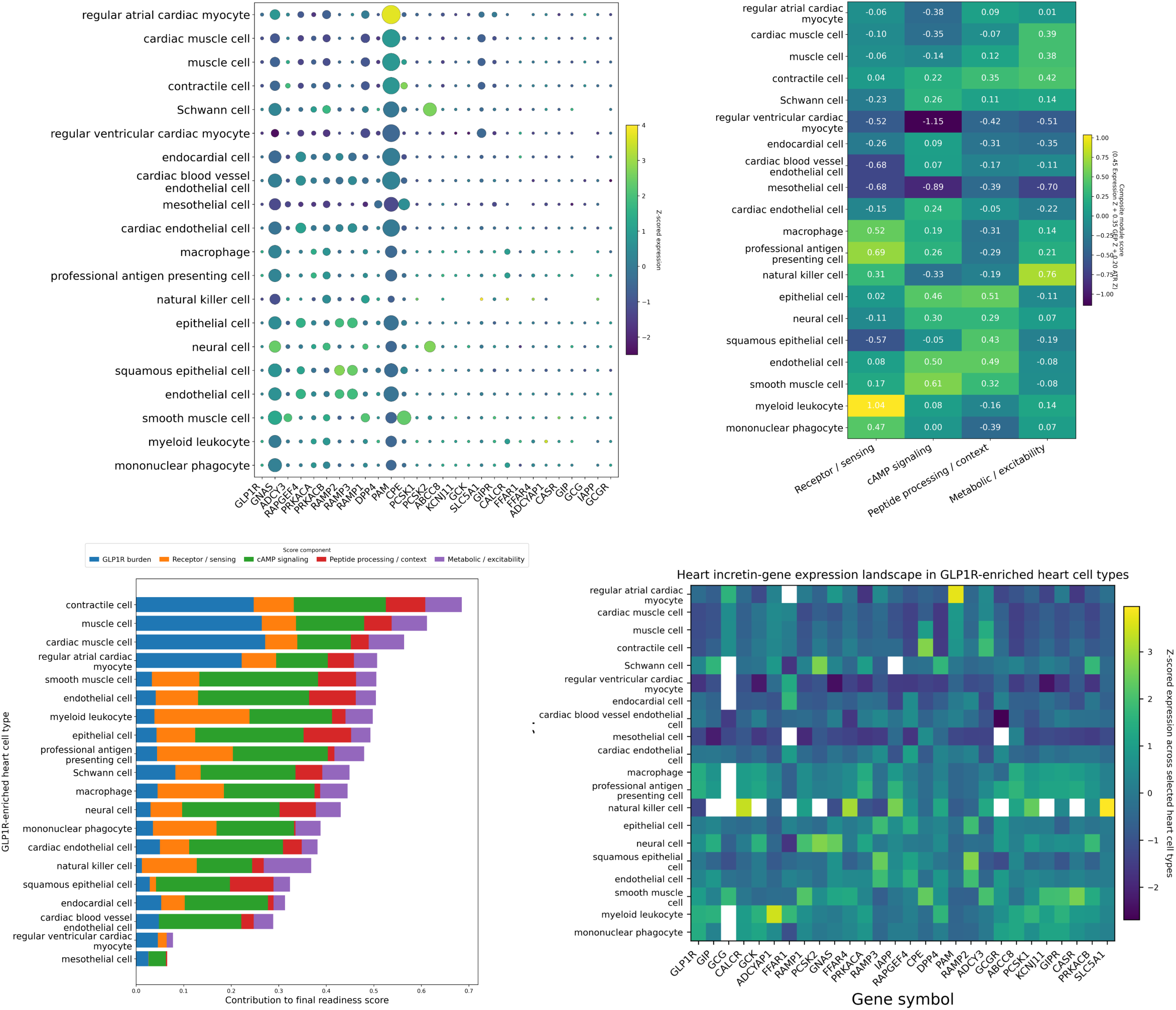
System of incretin genes and their expression patterns in single-cell RNA-sequencing data from heart tissue, with emphasis on GLP1R-enriched cardiovascular cell types. (Top left), Dot plot of semaglutide-relevant genes across GLP1R-enriched heart cell types. Dot size denotes the fraction of cells within each cell type expressing the indicated gene, and dot color denotes gene-wise z-scored expression across the selected heart cell types. **(Top Right),** Functional module heatmap summarizing semaglutide-response machinery across the same GLP1R-enriched heart cell types. Module scores represent composite z-scored signals derived from receptor and sensing genes, cAMP signaling genes, peptide-processing and contextual-modulation genes, and metabolic or excitability-associated genes. **(Bottom Left),** Ranked stacked-bar plot of a composite semaglutide functional-readiness score across GLP1R-enriched heart cell types. Total bar length represents the final readiness score, and colored segments denote the relative contribution from GLP1R burden, receptor and sensing context, cAMP signaling, peptide-processing context, and metabolic or excitability-associated programs. **(Bottom Right),** Heatmap of gene-wise z-scored expression across GLP1R-enriched heart cell types, highlighting coordinated enrichment patterns across cardiomyocyte-contractile, endothelial-epithelial, neuroglial and immune compartments. Together, these panels move beyond receptor detection alone to identify heart cell types that are transcriptionally positioned for potential semaglutide responsiveness.

**Table S1.** Clinical evidence of cardiovascular benefits for Semaglutide and Tizepatide.

| Drug | Population / condition | Cardiovascular findings established or supported in humans | Evidence tier | Key reference(s) |
| --- | --- | --- | --- | --- |
| Semaglutide | Overweight/obesity with established ASCVD, without diabetes | Reduced 3-point Major Adverse Cardiovascular Events (MACE), defined as cardiovascular death, nonfatal myocardial infarction, or nonfatal stroke | Randomized trial; label-supporting | SELECT / NEJM (New England Journal of Medicine) |
| Semaglutide | Type 2 diabetes with high cardiovascular risk | Reduced 3-point MACE versus placebo; strongest individual component signal was for nonfatal stroke; cardiovascular death alone was not significantly reduced | Randomized trial | SUSTAIN-6 / NEJM (New England Journal of Medicine) |
| Semaglutide | Obesity-related HFpEF | Improved heart-failure symptoms, physical limitations, exercise function, body weight, inflammatory measures and NT-proBNP; pooled analyses also showed reduction in cardiovascular death or worsening heart-failure events, driven mainly by fewer worsening heart-failure events | Randomized trials / pooled randomized analyses | STEP-HFpEF / NEJM, Circulation, JACC (New England Journal of Medicine) |
| Semaglutide | Obesity/overweight with prevalent heart failure in SELECT | Improved MACE and a heart-failure composite endpoint; favorable signals for cardiovascular death and all-cause death in the prevalent-heart-failure subgroup | Prespecified randomized subgroup analysis | Deanfield et al. / Lancet (The Lancet) |
| Semaglutide | ASCVD plus overweight/obesity, without diabetes | Associated with lower revised MACE-3, revised MACE-5, all-cause mortality, cardiovascular-related mortality and hospitalization for heart failure | Real-world observational | Smolderen et al. / Diabetes Obes Metab, JAHA/JACC-type reports (PMC) |
| Semaglutide | Broad clinical-practice comparator analyses | Associated with lower myocardial infarction or stroke composite risk versus sitagliptin-like comparator strategies in routine care | Real-world observational | Clinical-practice comparative analyses (PMC) |
| Tirzepatide | Obesity-related HFpEF | Reduced the composite of cardiovascular death or worsening heart-failure events, largely through fewer worsening heart-failure events; improved health status/KCCQ and exercise tolerance; cardiovascular death alone was not significantly reduced | Randomized trial | SUMMIT / NEJM, ACC, Circulation (New England Journal of Medicine) |
| Tirzepatide | Type 2 diabetes with established ASCVD | Demonstrated cardiovascular noninferiority versus dulaglutide for the composite of cardiovascular death, myocardial infarction, or stroke; superiority for MACE was not established | Randomized CV outcomes trial | SURPASS-CVOT / NEJM, ACC (American College of Cardiology) |
| Tirzepatide | Type 2 diabetes, BMI ≥25, pre-existing ischemic heart disease | Associated with lower risk of a composite of acute myocardial infarction, ischemic stroke and all-cause mortality; acute myocardial infarction and all-cause mortality were individually lower | Real-world observational | Dani et al. / JACC Advances-like observational study (JACC) |
| Tirzepatide | Broader real-world comparative analyses | Cardiovascular benefit in practice appears broadly comparable to semaglutide overall, with favorable but not definitive signals relative to active comparators | Real-world observational / comparative effectiveness | Observational comparative analyses and reviews (Taylor & Francis Online) |

**Table S2.** Baseline Demographic and Clinical Characteristics of Semaglutide-Treated Patients by Maximum Dose Reached During the 2-Year Observation Period.

| Dosage | Number of unique patients | Age, Mean (SD) | Female, % | T2DM Prevalence at Baseline, % | Baseline HBA1C, Mean (SD) |
| --- | --- | --- | --- | --- | --- |
| 0.25 mg | 2,192 | 57.1 (12.6) | 71.3 | 31.2 | 6.0 (1.3) |
| 0.5 mg | 3,383 | 60.1 (12.3) | 65.3 | 33.4 | 6.6 (1.7) |
| 1.0 mg | 2,386 | 59.9 (12.0) | 63.1 | 40.7 | 6.9 (1.7) |
| 1.7 mg | 353 | 53.4 (11.5) | 75.8 | 46.2 | 5.7 (0.5) |
| 2.0 mg | 1,095 | 59.4 (11.7) | 64.4 | 46.2 | 6.9 (1.7) |
| 2.4 mg | 1,108 | 53.6 (10.9) | 67.5 | 44.9 | 5.6 (0.6) |

**Table S3.** Baseline Demographic and Clinical Characteristics of Low-Dose and High-Dose Semaglutide-Treated Patients During the 2-Year Observation Period.

| Dose group | Number of unique patients | Age, Mean (SD) | Female, % | Baseline T2DM Prevalence, % | Baseline HBA1C, Mean (SD) | Number of Semaglutide prescriptions, Median [IQR] |  | Change in Semaglutide prescriptions (Post - Pre landmark), Median [IQR] |
| --- | --- | --- | --- | --- | --- | --- | --- | --- |
|  |  |  |  |  |  | 0 to 2 years (Landmark period) | 2-4 years (Post-landmark) |  |
| Low Dose (0.25mg - 1mg) | 8,725 | 59.3 (12.3) | 65.9 | 30.2 | 6.6 (1.7) | 1 [1-2] | 0 [0-2] | -1 [-2, -1] |
| High Dose (1.7mg+) | 3,794 | 57.8 (12.1) | 66.6 | 27.1 | 6.4 (1.5) | 5 [1-9] | 1 [0-2] | -4 [-9, -1] |

**Table S4.** Semaglutide prescription counts by weight-loss category before and after key treatment landmarks, median [IQR].

| Weight loss Category | Number of unique patients | Number of Semaglutide prescriptions, Median [IQR] |  |  | Change in Semaglutide prescriptions (Post - Pre landmark), Median [IQR] |
| --- | --- | --- | --- | --- | --- |
|  |  | 0 to 2 years (Landmark period) | 2-4 years (Post landmark) | (2 years post max weight loss attainment) |  |
| <5% | 4,050 | 1 [1, 2] | 0 [0, 2] | 0 [0, 2] | -1 [-3, -1] |
| 5-10% | 2,805 | 2 [1, 5] | 0 [0, 2] | 0 [0, 2] | -2 [-5, -1] |
| 10-15% | 1,943 | 2 [1, 7] | 0 [0, 2] | 1 [0, 2] | -3 [-7, -1] |
| 15-20% | 1,223 | 4 [1, 9] | 1 [0, 2] | 1 [0, 2] | -4 [-8, -1] |
| 20-25% | 707 | 5 [1, 10] | 1 [0, 2] | 1 [0, 2] | -5 [-9, -1] |
| 25%+ | 884 | 5 [2, 11] | 0 [0, 1] | 0 [0, 1] | -4 [-8, -1] |

**Table S5.**
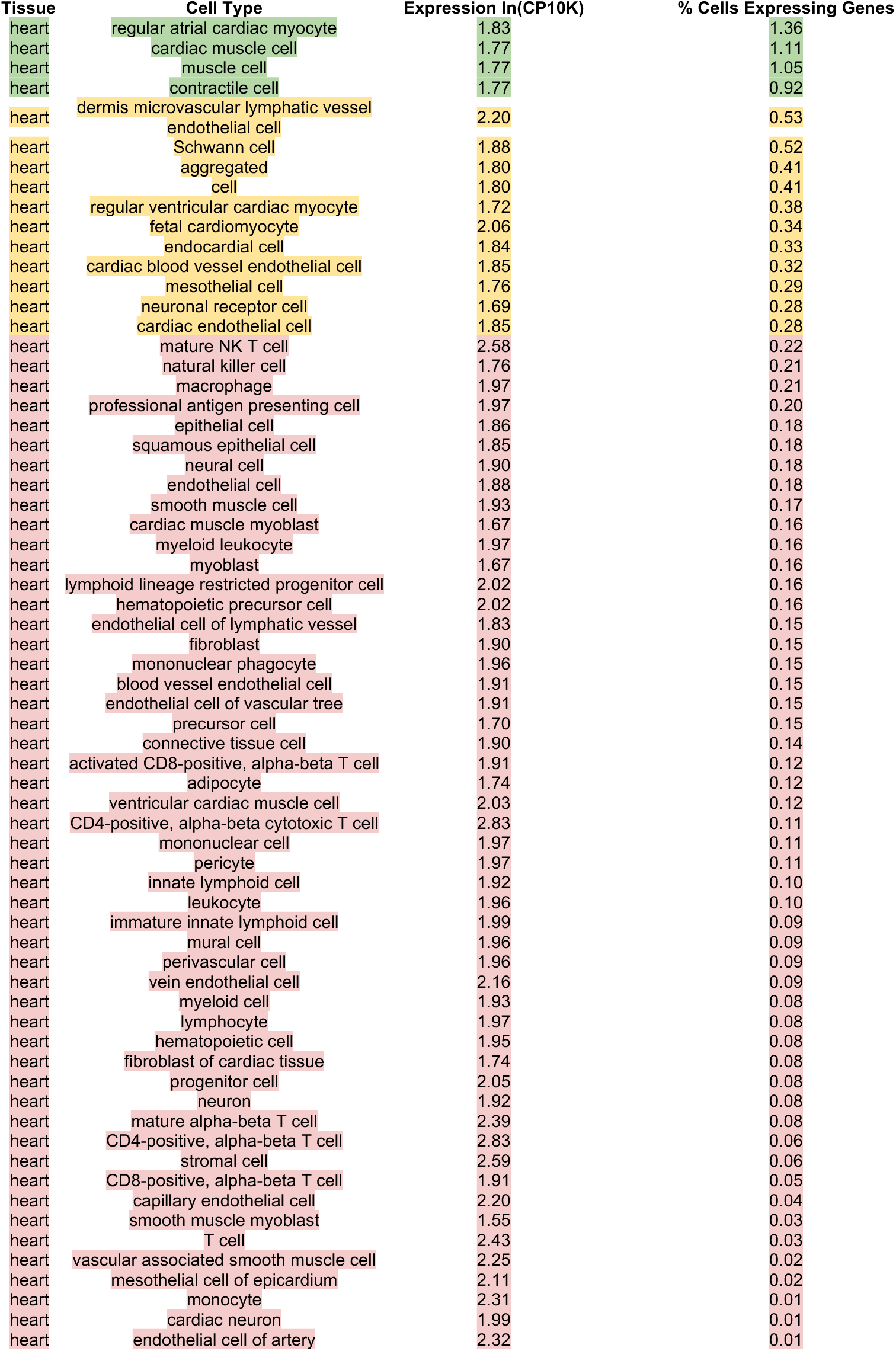
Single Cell RNA profile of GLP1R in the human heart.

**Table S6.** Constituent medications within the DPP-4 and SGLT2 comparator groups.

| **Comparator Group** | **Constituent Drugs** |
| --- | --- |
| DPP-4 inhibitors | sitagliptin, saxagliptin, linagliptin, alogliptin |
| SGLT2 inhibitors | empagliflozin, dapagliflozin, canagliflozin |

**Table S7.** ICD-9 and ICD-10 code definitions for baseline cardiovascular burden phenotypes used in subgroup assignment.

| **Phenotype** | **Disease Name** | **ICD10 Code Prefixes** | **ICD9 Code Prefixes** |
| --- | --- | --- | --- |
| Ischemic heart disease | Ischemic heart disease | I20, I21, I22, I23, I24, I25 | 410, 411, 412, 413, 414 |
| Cerebrovascular disease | Cerebrovascular disease | I60, I61, I62, I63, I64, I65, I66, I67, I68, I69 | 430, 431, 432, 433, 434, 435, 436, 437, 438 |
| Hypertension | Hypertension | I10, I11, I12, I13, I14, I15, I16 | 401, 402, 403, 404, 405 |
| Heart failure | Heart failure | I50 | 428 |
| Arrhythmia / Conduction disorders | Arrhythmias / conduction disorders | I44, I45, I46, I47, I48, I49 | 426, 427 |
| Peripheral / Vascular disease atherosclerosis | Peripheral vascular disease / atherosclerosis | I70, I73, I74 | 440, 443, 444 |
| Valvular / rheumatic heart disease | Valvular / rheumatic heart disease | I00, I01, I02, I05, I06, I07, I08, I09, I34, I35, I36, I37, I38, I39 | 390, 391, 392, 393, 394, 395, 396, 397, 398, 424 |
| Cardiomyopathy | Cardiomyopathy | I42, I43 | 425 |
| Aortic disease | Aortic disease | I71 | 441 |
| Venous thromboembolism | Venous thromboembolism | I26, I80, I81, I82 | 415.1, 451, 452, 453 |

**Table S8.** Interval between index date and most recent qualifying preindex CV diagnosis.

| Cohort | Cardiovascular subgroup | Number of unique patients | Gap to most recent qualifying preindex ICD, median [IQR], d |
| --- | --- | --- | --- |
| semaglutide | all cv conditions | 53114 | 108 [26, 340] |
| semaglutide | aortic disease | 1013 | 416 [165, 740] |
| semaglutide | arrhythmia conduction disorder | 10611 | 214 [49, 639] |
| semaglutide | cardiomyopathy | 1735 | 197 [50, 672] |
| semaglutide | cerebrovascular disease | 3147 | 359 [111, 987] |
| semaglutide | heart failure | 4221 | 137 [36, 431] |
| semaglutide | hypertension | 45704 | 118 [30, 344] |
| semaglutide | ischemic heart disease | 10206 | 162 [43, 443] |
| semaglutide | peripheral vascular disease atherosclerosis | 3745 | 238 [77, 679] |
| semaglutide | valvular rheumatic heart disease | 3441 | 315 [92, 831] |
| semaglutide | venous thromboembolism | 3122 | 608 [159, 1,535] |
| metformin | all cv conditions | 132970 | 55 [6, 264] |
| metformin | aortic disease | 2911 | 225 [44, 666] |
| metformin | arrhythmia conduction disorder | 25457 | 139 [13, 590] |
| metformin | cardiomyopathy | 3864 | 145 [22, 579] |
| metformin | cerebrovascular disease | 10332 | 239 [38, 831] |
| metformin | heart failure | 9175 | 81 [9, 374] |
| metformin | hypertension | 109617 | 63 [7, 260] |
| metformin | ischemic heart disease | 30177 | 128 [15, 496] |
| metformin | peripheral vascular disease atherosclerosis | 9057 | 163 [25, 555] |
| metformin | valvular rheumatic heart disease | 8439 | 263 [40, 824] |
| metformin | venous thromboembolism | 6261 | 346 [41, 1,170] |
| DPP-4 inhibitors | all cv conditions | 11875 | 88 [11, 398] |
| DPP-4 inhibitors | aortic disease | 298 | 282 [80, 764] |
| DPP-4 inhibitors | arrhythmia conduction disorder | 3105 | 132 [17, 476] |
| DPP-4 inhibitors | cardiomyopathy | 632 | 181 [32, 588] |
| DPP-4 inhibitors | cerebrovascular disease | 1321 | 233 [54, 808] |
| DPP-4 inhibitors | heart failure | 2217 | 75 [11, 345] |
| DPP-4 inhibitors | hypertension | 9558 | 113 [19, 437] |
| DPP-4 inhibitors | ischemic heart disease | 3838 | 151 [26, 568] |
| DPP-4 inhibitors | peripheral vascular disease atherosclerosis | 1471 | 165 [39, 533] |
| DPP-4 inhibitors | valvular rheumatic heart disease | 1221 | 224 [39, 655] |
| DPP-4 inhibitors | venous thromboembolism | 665 | 298 [36, 1,006] |
| SGLT2 inhibitors | all cv conditions | 74169 | 24 [4, 128] |
| SGLT2 inhibitors | aortic disease | 3577 | 308 [84, 621] |
| SGLT2 inhibitors | arrhythmia conduction disorder | 33798 | 42 [7, 220] |
| SGLT2 inhibitors | cardiomyopathy | 14410 | 55 [9, 259] |
| SGLT2 inhibitors | cerebrovascular disease | 8543 | 252 [55, 812] |
| SGLT2 inhibitors | heart failure | 34279 | 21 [4, 105] |
| SGLT2 inhibitors | hypertension | 56957 | 65 [11, 249] |
| SGLT2 inhibitors | ischemic heart disease | 31601 | 59 [10, 251] |
| SGLT2 inhibitors | peripheral vascular disease atherosclerosis | 10912 | 154 [35, 468] |
| SGLT2 inhibitors | valvular rheumatic heart disease | 16479 | 101 [15, 394] |
| SGLT2 inhibitors | venous thromboembolism | 4748 | 410 [71, 1,323] |

**Table S9.** ICD-9 and ICD-10 code definitions for the composite cardiovascular endpoint, including fatal and nonfatal components.

| Component | ICD10 Code Prefixes | ICD9 Code Prefixes |
| --- | --- | --- |
| All-cause death | N/A | N/A |
| Myocardial infarction/Acute coronary syndrome | I21, I22, I23, I24 | 410, 411 |
| Stroke/cerebrovascular event | I60, I61, I62, I63, I64, I65, I66, I67, I68, I69 | 430, 431, 432, 433, 434, 435, 436, 437, 438 |

## Supplementary Text

### Cardiac GLP1R geography provides biologic plausibility

The expression data provide a plausible biological framework for this clinical pattern. Prior work has shown that GLP1R is detectable in human cardiac tissue and that cardiac or endocardial/endothelial GLP-1R signaling can mediate cardioprotective effects in experimental systems^7,19^. Our atlas-based analyses extend that rationale in two ways. First, they show that the human heart is not transcriptionally silent for GLP1R at the tissue level. Second, they resolve that signal into distinct cell populations.

Following is a potential dose-stratified interpretation of the cardiac GLP1R atlas. At lower semaglutide exposure, pharmacologic activity may be concentrated in the most prevalent GLP1R-expressing cardiac populations, particularly atrial and broader cardiomyocyte, or contractile-cell compartments, in which GLP1R-positive cells are relatively more frequent (**Table S5**, green-highlighted rows). At moderate to high exposure, engagement may extend to a broader set of less prevalent but still recurrent endothelial, endocardial, vascular-associated, and stromal populations (**Table S5**, yellow-highlighted rows), potentially broadening favorable effects on vascular tone, myocardial loading conditions, endothelial biology and inflammatory signaling. At very high exposure, semaglutide may additionally engage rare GLP1R-expressing immune-cell subsets, including CD4-positive alpha-beta T-cell and related lymphoid compartments (**Table S5**, red-highlighted rows).

### Whole-body receptor geography and implications for organ-directed pharmacology

The whole-body single-cell atlas further refines this interpretation by showing that GLP1R tissue geography depends on how receptor burden is summarized. Prevalence-weighted GLP1R Engagement Potential (GEP) prioritizes tissues in which GLP1R-positive cells are comparatively frequent, whereas Absolute Target Load (ATL) prioritizes tissues containing the largest aggregate reservoir of GLP1R-positive cells. In this framework, systemic metabolic effects and organ-specific cardiovascular effects need not arise from the same receptor geography. Rather, semaglutide exposure may simultaneously engage pancreatic and other whole-body compartments that shape weight loss while also interacting with a substantial cardiac target reservoir capable of supporting direct heart-specific biology.

### Distributed GLP1R signaling programs across myocardial, vascular, cardio-immune cells

Examination of GLP1R signal in the heart shows distribution across several biologically meaningful compartments rather than being restricted to a single rare niche (**Fig. S6 - top left panel**). Cardiomyocyte-contractile states, especially regular atrial cardiac myocytes, cardiac muscle cells, muscle cells and contractile cells, show the strongest overall GLP1R burden, consistent with the idea that the myocardium itself is a major transcript-level reservoir of semaglutide responsiveness. At the same time, the data shows that these cardiomyocyte-like compartments also co-express canonical downstream signaling genes of GLP1R such as GNAS, ADCY3, RAPGEF4, PRKACA and PRKACB, suggesting that receptor detection is embedded within a productive signaling framework rather than occurring in isolation (**Fig. S6 - top left panel**). Vascular-interface states, including endothelial, cardiac endothelial, endocardial and cardiac blood vessel endothelial cells, show recurrent enrichment of RAMP2, RAMP3, RAPGEF4, DPP4 and related contextual genes, a pattern that is concordant with literature implicating GLP-1 receptor agonists in endothelial protection, vascular anti-inflammatory signaling and attenuation of atherosclerotic biology^4–7^. Finally, myeloid, macrophage and antigen-presenting populations show distinct enrichment of CALCR, ADCYAP1, FFAR1 and related signaling genes, raising the possibility that part of semaglutide’s cardiovascular benefit may be mediated through immunometabolic remodeling rather than through cardiomyocytes alone, a concept that is broadly compatible with the known anti-inflammatory and cardiometabolic effects of GLP-1 receptor agonism^4,6,7^.

### Computing potential for functional semaglutide-response modules

We computed a heatmap to distill this gene-level complexity into a functional architecture (**Fig. S6 - top right panel**). Cardiomyocyte-contractile compartments retain comparatively strong receptor burden and appreciable cAMP signaling competence, supporting the interpretation that semaglutide-responsive biology in heart may include direct myocardial interfaces. In contrast, endothelial and epithelial compartments are especially notable for stronger peptide-processing or contextual-modulation signatures, including RAMP2, RAMP3, DPP4, CPE and RAPGEF4, suggesting that these cell types may be particularly important for vascular and tissue-interface responses to GLP-1 receptor agonism (**Fig. S6 - top right panel**). Myeloid leukocytes, macrophages and professional antigen-presenting cells show comparatively prominent receptor and sensing module scores, which is particularly interesting because semaglutide has reduced major adverse cardiovascular events in people with overweight or obesity and established cardiovascular disease in the SELECT trial, while related work increasingly points to inflammatory and vascular mechanisms as important mediators of benefit, beyond weight loss alone^1,4,6,7^. The relatively distinct Schwann-cell and neural-cell module patterns suggest a peripheral neuroglial niche that may participate in incretin biology, but this remains more hypothesis-generating than the vascular-inflammatory axis, for which the clinical and experimental literature is already stronger^4,6,7^.

### Towards a composite functional-readiness framework for semaglutide cardiac activity

The functional-readiness ranking is useful because it prioritizes cell types not simply by GLP1R abundance but by the combination of receptor burden, receptor-context genes, downstream cAMP machinery, peptide-processing capacity and metabolic or excitability-associated effectors (**Fig. S6 - bottom left panel**). In this framework, contractile and muscle-like states rank highly because they combine substantial GLP1R burden with signaling genes such as GNAS, ADCY3 and RAPGEF4 and with downstream contextual genes including CPE and PAM. Smooth-muscle and endothelial-interface states also emerge as highly ranked, which is biologically plausible given the substantial literature on GLP-1 receptor agonist effects on endothelial function, vascular inflammation and atherosclerotic remodeling^6,7^. Myeloid leukocytes, macrophages and antigen-presenting cells rank prominently because they combine receptor-adjacent sensing programs with inflammatory and peptide-context genes, supporting an immunometabolic interpretation of semaglutide action that is consistent with the cardiovascular and HFpEF clinical data (**Fig. S6 - bottom left panel**). In SELECT, semaglutide lowered cardiovascular event risk in overweight or obesity without diabetes, and in STEP-HFpEF and STEP-HFpEF DM it improved symptoms, physical limitations and exercise-related outcomes in obesity-related HFpEF, indicating that the net cardiovascular phenotype of semaglutide likely reflects integrated myocardial, vascular and inflammatory effects rather than a single-cell-type mechanism^1–4^.

### Single cell RNA patterns hint at broad model of possible semaglutide cardiac activity

The expression heatmap provides the gene-level resolution underlying the higher-order summaries (**Fig. S6 - bottom right panel**). Several striking patterns stand out. First, cardiomyocyte-contractile states show coordinated expression of GLP1R with GNAS, CPE, PAM, RAPGEF4 and ADCY3, supporting a model in which receptor-positive myocardial compartments may be capable of signal transduction rather than merely harboring sparse receptor transcripts. Second, endothelial, epithelial and squamous epithelial states show recurrent RAMP2, RAMP3, RAPGEF4 and DPP4 signals, consistent with an interface biology centered on peptide sensing and vascular or barrier regulation (**Fig. S6 - bottom right panel**). Third, immune and myeloid compartments show notable enrichment of CALCR, ADCYAP1 and FFAR1, suggesting that incretin-adjacent and nutrient-sensing programs may intersect in inflammatory cell states relevant to cardiovascular remodeling. Finally, neural and Schwann compartments show PCSK2-rich signatures, implying specialized neuropeptide-processing capacity. The overall picture is that semaglutide activity potential in heart is most plausibly distributed across three major axes: a myocardial-contractile axis, a vascular-interface axis and an inflammatory-immunometabolic axis, with a smaller neuroglial niche that deserves follow-up. This distributed model is more congruent with current clinical and mechanistic literature than a narrow interpretation based on cardiomyocytes or endothelial cells alone^4–6^.

### Implications for endpoint selection, dose optimization and high-dose extrapolation

The most defensible interpretation of our cumulative single cell RNA analysis is that semaglutide’s cardiovascular benefit is unlikely to be explained solely by receptor presence in one dominant heart cell type. Instead, the data support a distributed-response model in which GLP1R-positive cardiomyocyte-contractile cells provide one plausible interface, endothelial and endocardial cells provide a strong vascular-context interface, and macrophage-myeloid populations provide an inflammatory-immunometabolic interface. This interpretation is aligned with the clinical observation that semaglutide reduces cardiovascular events in obesity without diabetes and improves obesity-related HFpEF outcomes, and with mechanistic literature emphasizing vascular protection, reduced endothelial dysfunction and attenuation of inflammatory remodeling as important components of GLP-1 receptor agonist biology^20–24^.

Although weight reduction likely contributes to benefit for selected outcomes, the absence of a consistent monotonic association between achieved weight loss and cardiovascular risk suggests additional organ-directed mechanisms. Semaglutide’s cardiovascular effects may therefore reflect not only systemic changes in adiposity, but also direct or indirect actions on cardiac, vascular, and immune biology, including effects on glycemic physiology, blood pressure, vascular inflammation, endothelial function, myocardial loading conditions, neurohormonal signaling and arrhythmic susceptibility.

These findings have immediate translational implications. Current incretin development and clinical use are often benchmarked primarily against the magnitude of total-body weight reduction. Our results argue that this strategy is too narrow for cardiovascular medicine. In the present framework, weight loss is a whole-body pharmacodynamic readout, but cardiovascular protection may also reflect organ-directed biology. That distinction matters for dose optimization, endpoint selection and cross-drug comparison. More broadly, these data support incorporating dedicated cardiovascular endpoints, and not weight loss alone, into therapeutic optimization.

Extrapolation of this empirical real-world relationship to semaglutide 7.2 mg yields a hypothetical maximum weight change of approximately −29.8% over the 2-year landmark period (**Fig. 2**). While our analysis is based on maximum dose reached and the lowest observed pre-landmark weight over 2 years in routine care, the STEP UP trial evaluated a protocolized once-weekly 7.2 mg semaglutide high-dose regimen over 72 weeks. These clinical trial outcomes appear directionally concordant, with STEP UP showing approximately 20.7% mean loss under the efficacy estimate and 31.2% of participants achieving at least 25% weight loss^21^. Our landmark-derived, real-world model based on maximum attained dose and lowest observed pre-landmark weight is closer to an upper-envelope estimate of achievable pharmacologic effect than to a trial-average endpoint measured at a fixed week under protocolized titration, retention, and adherence conditions. In that context, the extrapolated value of ∼29.8% understandably sits above the trial-average response but within the broader range suggested by the high-responder tail of STEP UP. The implication is that real-world pharmacological imputation can be useful for estimating directionality and attainable response range at higher exposures but should not be treated as a one-to-one surrogate for randomized-trial mean efficacy when dose-escalation schedules, treatment persistence, patient selection and endpoint definitions differ.

